# Genomic and Transcriptomic Determinants of Resistance to CDK4/6 Inhibitors and Response to Combined Exemestane plus Everolimus and Palbociclib in Patients with Metastatic Hormone Receptor Positive Breast Cancer

**DOI:** 10.1101/2022.07.11.22277416

**Authors:** Jorge Gómez Tejeda Zañudo, Romualdo Barroso-Sousa, Esha Jain, Qingchun Jin, Tianyu Li, Jorge E. Buendia-Buendia, Alyssa Pereslete, Daniel L. Abravanel, Arlindo R. Ferreira, Eileen Wrabel, Karla Helvie, Melissa E. Hughes, Ann H. Partridge, Beth Overmoyer, Nancy U. Lin, Nabihah Tayob, Sara M. Tolaney, Nikhil Wagle

**Author notes:** E. Jain.: Repare Therapeutics, Cambridge, Massachusetts, USA. Current address for J. E. Buendia-Buendia: Cellarity, Somerville, Massachusetts, USA. Current address for A. R. Ferreira: Breast Unit, Champalimaud Clinical Center, Champalimaud Foundation, Portugal. J. Gómez Tejeda Zañudo and R. Barroso-Sousa contributed as co-first authors of this article. S. M. Tolaney and N. Wagle contributed as co-senior authors of this article. Corresponding Authors: S. M. Tolaney,; N. Wagle,.

## Abstract

Even though multiple resistance mechanisms and pathways for cyclin-dependent kinase 4/6 inhibitors (CDK4/6i) have been discovered, the complete landscape of resistance is still being elucidated. Moreover, the optimal subsequent therapy to overcome resistance remains uncertain.

To address this, we carried out a phase I/II clinical trial of exemestane plus everolimus and palbociclib, triplet therapy for CDK4/6i-resistant hormone receptor–positive (HR+), HER2-metastatic breast cancer, one of the first trials evaluating CDK4/6i after CDK4/6i progression. With an observed clinical benefit rate of 18.8% (*n* = 6/32), the trial did not meet its primary efficacy endpoint. However, we leveraged the multi-omics tumor data from these patients to study the landscape of CDK4/6i resistance and to identify correlates of response to triplet therapy.

We generated whole exome sequencing from 24 tumor and 17 ctDNA samples and transcriptome sequencing from 27 tumor samples obtained from 26 patients in the trial. Genomic and evolutionary analysis recapitulated the spectrum of known resistance genes (*ERBB2, NF1, AKT1, RB1, ESR1*) and pathways (RTK/MAPK, PI3K/AKT/mTOR, cell cycle, estrogen receptor), discovered potential new mechanisms of resistance in these pathways (*ERBB2* amplification, *BRAF*^V600E^, *MTOR*^T1977R^), and identified a patient with co-existing tumor lineages with distinct activating *ERBB2* mutations, potentially the first case of convergent evolution of HER2 activation following CDK4/6i therapy. Joint genomic and transcriptomic analysis revealed that genomic resistance mechanisms were associated with transcriptomic features in their respective pathways, suggesting that transcriptomic features could be used to identify the pathways driving resistance. In particular, the mutually exclusive *ESR1* and *ERBB2*/*BRAF* mutations, were each linked with high activity in distinct pathway signatures (estrogen receptor pathway vs RTK/MAPK pathway, respectively) and were exclusive to distinct molecular subtypes (Luminal A or Luminal B vs HER2-E, respectively). Overall, incorporating clinical and multi-omics features in CDK4/6i-resistant tumors enabled identification of known or putative drivers of resistance to the prior CDK4/6i and anti-estrogen therapies in nearly every patient (*n* = 22/23), including several patients in which transcriptomic features were the sole drivers. Genomic and transcriptomic features – particularly PI3K/AKT/mTOR mutations and/or high mTORC1 pathway activity - suggested that clinical benefit to combined estrogen receptor, CDK4/6, and mTOR inhibition was correlated with activation of the mTOR pathway.

Our results illustrate how transcriptome sequencing provides complementary and additional information to genome sequencing, and how integrating both may help better identify patients likely to respond to CDK4/6i therapies.

**Significance:** Combined endocrine, CDK4/6 inhibitor, and mTOR inhibitor therapy showed limited benefit in patients with HR+ metastatic breast cancer who had progressed on a prior CDK4/6 inhibitor. Multi-omics analysis of tumors from this trial identified novel genomic and transcriptomic drivers of CDK4/6i resistance, known or putative drivers of resistance in 22/23 patients, and correlates of response to the trial therapy. Integrated genome and transcriptome sequencing may better identify factors that determine response to CDK4/6i therapy and help select optimal therapy.

## Introduction

The combination of cyclin-dependent kinase 4/6 inhibitors (CDK4/6i) (palbociclib, ribociclib, abemaciclib) and endocrine therapy has become standard of care for the treatment of patients with hormone-receptor positive (HR+), human epidermal growth factor receptor 2 negative (HER2-) metastatic breast cancer (MBC). However, some patients do not respond at all to CDK4/6i therapy (intrinsic resistance) while other patients initially respond but eventually become refractory to therapy (acquired resistance), which results in disease progression.

Multiple studies in the last few years have started to reveal the genomic landscape of resistance to CDK4/6 inhibitors (Spring et al. 2020). One major class of resistance mechanisms that have been observed in the clinical setting are alterations that activate the cell cycle pathway, namely, loss-of-function alterations in *RB1* (Condorelli et al. 2018; O’Leary et al. 2018; Z. Li et al. 2018; S. A. Wander et al. 2020) and high cyclin E1 (*CCNE1*) mRNA expression (N. C. Turner et al. 2019). Other notable potential resistance mechanisms in the cell cycle pathway are amplifications of *AURKA* (S. A. Wander et al. 2020) and CDK6 overexpression (Yang et al. 2017; Cornell et al. 2019; Q. Li et al. 2022).

In addition to the cell cycle pathway, multiple oncogenic pathways have been associated with resistance to CDK4/6 inhibitors in the clinical setting. In the estrogen receptor (ER) pathway, low ER expression (S. A. Wander et al. 2020; N. C. Turner et al. 2019) and basal molecular subtype (Aleix Prat et al. 2021) have been associated with a lack of response to CDK4/6 inhibitors. Loss-of-function of FAT1, which activates the Hippo pathway and results in an increase in *CDK6* expression, has been associated with poor response to CDK4/6i therapy (Z. Li et al. 2018). Oncogenic alterations in the receptor tyrosine kinase (RTK) (*FGFR1, FGFR2, ERBB2*), MAPK (*KRAS, HRAS, NRAS*), and PI3K/AKT/mTOR (*AKT1, PTEN*) pathways, all of which are upstream of the cell cycle pathway, have been found to be enriched in patient samples that are resistant to CDK4/6 inhibitors and to cause resistance in pre-clinical models (Nayar et al. 2019; Mao et al. 2020; S. A. Wander et al. 2020; Costa et al. 2020; Formisano et al. 2019; N. Turner et al. 2010).

Despite the variety of genomic resistance mechanisms and pathways identified by this recent work, the complete landscape of CDK4/6i resistance is still being elucidated. In particular, the transcriptomic landscape has remained mostly unexplored.

Oncogenic alterations in the RTK, MAPK, and PI3K/AKT/mTOR pathway have also been found to confer resistance to endocrine therapies (Razavi et al. 2018; Bertucci et al. 2019; Nayar et al. 2019; Mao et al. 2020; Hanker, Sudhan, and Arteaga 2020). This is analogous to the role that alterations in these pathways play in CDK4/6 therapy, since the canonical resistance mechanisms to endocrine therapy are alterations that directly affect the estrogen receptor (primarily activating ESR1 mutations) (Razavi et al. 2018; Bertucci et al. 2019; Hanker, Sudhan, and Arteaga 2020).

Given that RTK, MAPK, and PI3K/AKT/mTOR oncogenic alterations drive resistance to both endocrine therapy and CDK4/6i,targeting these pathways might be able to overcome resistance. For endocrine-resistant MBC, the BOLERO-2 phase III trial found that the combination of exemestane (an aromatase inhibitor) and everolimus (mTOR inhibitor) resulted in a longer progression-free survival than exemestane alone (Baselga et al. 2012). Whether an analogous result holds for the combination of CDK4/6i, endocrine therapy, and everolimus or other PI3K/AKT/mTOR targeted therapy is still an open question for CDK4/6i-resistant MBC.

Here we report the final results from a phase I/II clinical trial evaluating the safety and efficacy of exemestane plus everolimus and palbociclib, triplet therapy in patients with CDK4/6i-resistant and endocrine-resistant HR+ MBC (NCT02871791). By leveraging multi-omics data from patients that participated in this trial – whole-exome sequencing (WES) and RNA sequencing (RNA-seq) of tumor biopsies and circulating tumor DNA (ctDNA), accompanied by comprehensive clinical data – we performed an integrated analysis of the molecular correlates of both the landscape of resistance to initial CDK4/6 inhibitors and the response to triplet therapy (Fig. 1A). Our analysis identified genomic alterations in resistance genes (*ESR1, ERBB2, AKT1, RB1, MTOR, BRAF*), genomic/transcriptomic resistance features (concurrent HER2-enriched molecular subtype and RTK/MAPK oncogenic mutations, concurrent ER pathway activity and *ESR1* oncogenic mutations), evidence of convergent evolution of *ERBB2* activation following progression on CDK4/6i therapy, and indicators that the mTOR pathway activity is associated with response to triplet therapy. The results are an important step towards elucidating the complete landscape of resistance to CDK4/6i in HR+/HER2− MBC and highlight how multi-omics data may better identify factors that determine response to CDK4/6i therapy.

**Fig. 1.**
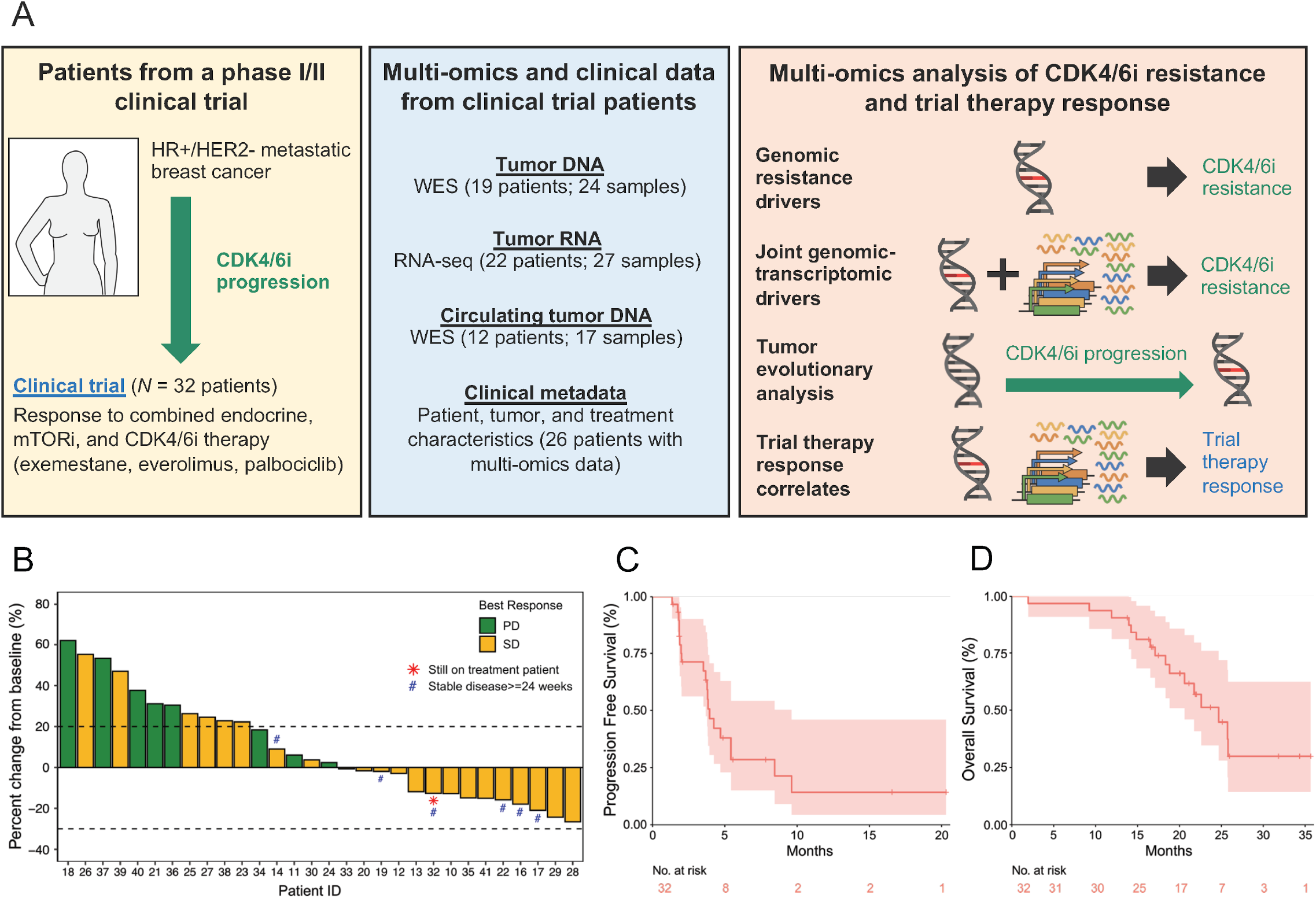
Methodological overview of this work and plots for the phase II portion of the clinical trial. (A) Graphical methodological overview. (B) Waterfall plot of best percentage change from baseline of tumor lesions. Patients 15 and 31, who stopped treatment before tumor response could be evaluated, were excluded from this plot. (C) Progression-free survival (PFS) Kaplan-Meier curve. The median PFS is 3.94 months (95% CI: 3.68-9.63). (D) Overall survival (OS) Kaplan-Meier curve. The median OS is 24.7 months (95% CI: 20.6 – N/A). *n* = 32 patients were included in the phase II portion of the trial.

## Results

### Patients received limited clinical benefit from triplet therapy

Patients participated in a phase I/II clinical trial (NCT02871791) evaluating the safety and efficacy of triplet therapy: palbociclib (a CDK4/6 inhibitor) + everolimus (mTOR inhibitor) + exemestane (steroidal aromatase inhibitor). Eligible patients had been diagnosed with HR+/HER2− MBC and had progressed on a prior CDK4/6i and a prior endocrine therapy (a nonsteroidal aromatase inhibitor).

For the phase I portion of the trial, 9 patients were enrolled. The recommended phase 2 dose (RP2D) was 100 mg palbociclib + 5 mg everolimus + 25 mg exemestane, with a MTD of 100 mg for palbociclib. For the phase II portion of the trial, 32 patients were enrolled (Supplemental Table S1). All patients were female and the median age (range) was 55.5 years (36-73) (Supplemental Table S2). Approximately one-third of patients had received one line of prior chemotherapy in the metastatic setting (*n* = 12/32, 37.5%), the remainder had received no prior chemotherapy (n = 20/32, 62.5%) (Supplemental Table S2). Almost all patients had received at least one prior line of endocrine therapy (*n* = 31/32, 96.9%), and over half had received 2 or more prior lines (*n* = 17/32, 53.1%). All but 2 patients had received palbociclib (*n* = 30/32, 93.8%) and the remaining 2 patients had received abemaciclib (*n* = 2/32, 6.3%). With a median follow-up (interquartile range) of 23.7 months (20.1-31.8 months), 6 patients had stable disease (SD) ≥ 24 weeks (*n* = 6/32, 18.8%), 18 patients had SD ≥ 12 weeks (*n* = 18/32, 56.3%), and 8 patients had progressive disease (PD) (*n* = 8/32, 25.0%) as best response (Fig. 1B, Supplemental Table S3). The clinical benefit rate (CBR, CR + PR + SD ≥ 24 weeks) was 18.8%, which was below the pre-specified efficacy endpoint of CBR ≥ 65%. Median progression-free survival (PFS) was 3.94 months (95% CI: 3.68-9.63) and median overall survival (OS) was 24.7 months (95% CI: 20.6 - not reached) (Fig. 1C, 1D). Additional information on the clinical trial can be found in the Supplemental Material, which includes a more detailed summary of the methods and results of the clinical trial (Supplemental Text), patient disposition at time of data cutoff (Supplemental Table S1), baseline patient, tumor, and treatment characteristics (Supplemental Table S2), best response by RECIST (Supplemental Table S3), adverse effects and dose reductions for the study drugs (Supplemental Table S4, Supplemental Table S5), and patient-level clinical characteristics (Supplemental Table S6).

### Whole exome and transcriptome sequencing of baseline tumor and ctDNA revealed known and novel resistance mechanisms to CDK4/6 inhibitors and endocrine therapy

We generated WES and RNA-seq data from tumor biopsies and ctDNA samples from patients who participated in the phase II portion of the clinical trial of triplet therapy (Fig. 2A, Supplemental Table S7, Supplemental Table S8). As part of the clinical trial, we collected a research tumor biopsy and blood sample at baseline (after progression on the prior CDK4/6 inhibitor but before initiation of triplet therapy) and additional serial blood samples while on the trial. Additionally, when possible, we acquired archival tumor biopsies that preceded the patient’s initial exposure to a CDK4/6i to serve as a CDK4/6i-naive sample. Given the exploratory nature of this study and our limited sample size, significance tests in our analyses were not corrected for multiple comparisons.

**Fig. 2.**
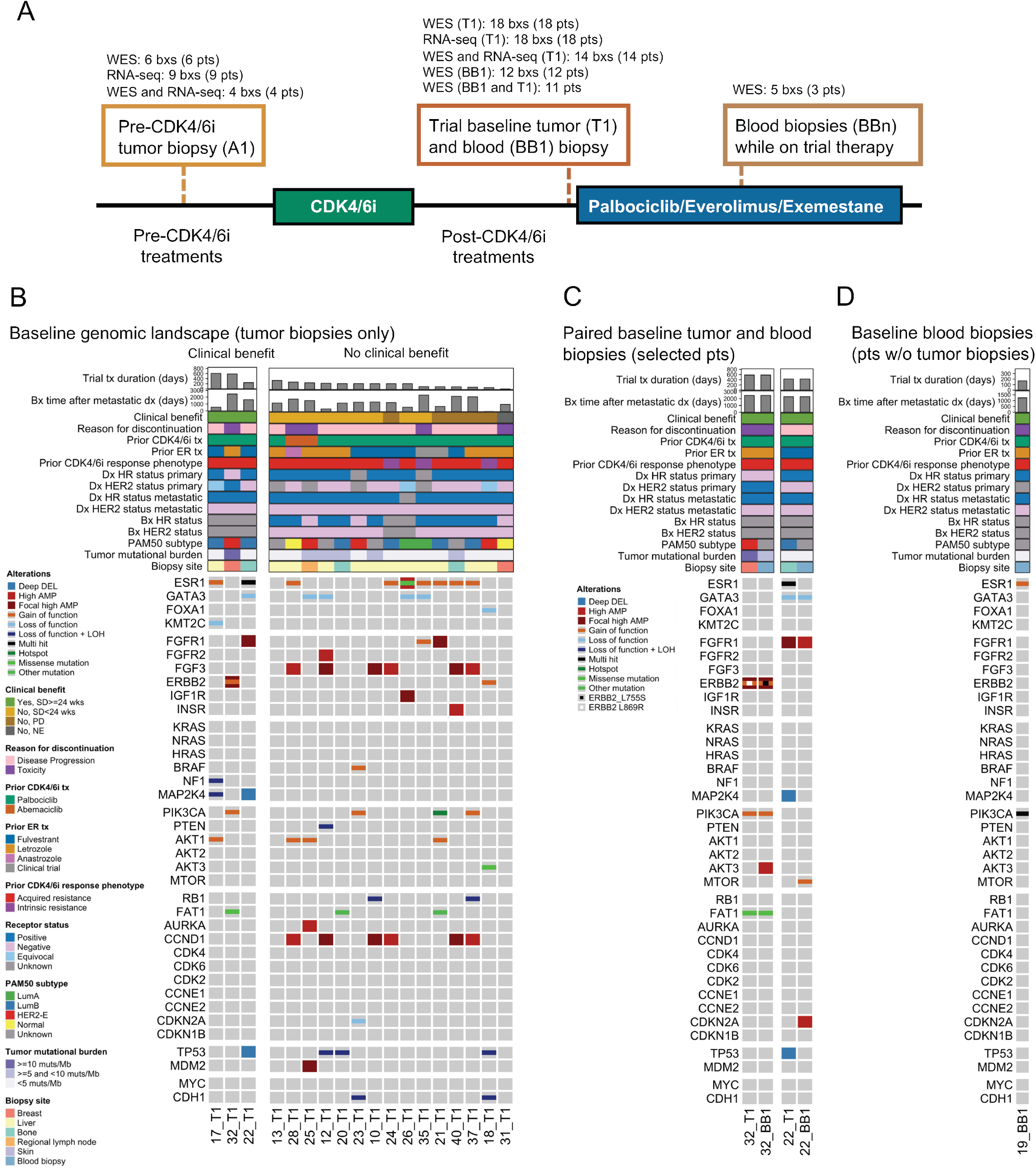
Genomic landscape of resistance to CDK4/6 inhibitors in clinical trial baseline biopsies. The genomic landscape recapitulates known driver genes and pathways of CDK4/6i resistance and putative driver genes and mutations (*BRAF*^V600E^, *MTOR*^T1977R^, *PIK3CA*^E545K,G1007R^). (A) Cohort of tumor and blood biopsies used for multi-omics analysis and their timing. Patients received triplet therapy (palbociclib + everolimus + exemestane) as part of the clinical trial, and had progressed on a prior CDK4/6i and a prior endocrine therapy. (B-D) Comutation plots (CoMut) representing the genomic landscape of baseline tumor and blood biopsies from the clinical trial. All baseline tumor biopsies are shown in panel B (*n* = 18 samples from *n* = 18 patients); paired baseline tumor and blood biopsies from patients with distinct co-existing tumor lineages are shown in panel C (*n* = 4 samples from *n* = 2 patients); baseline blood biopsies from patients with no paired tumor biopsy are shown in panel D (*n* = 1 sample from *n* = 1 patient). In each panel, biopsies are ordered by treatment duration on triplet therapy. Copy-number alterations and nonsynonymous mutations from selected genes (including all from (S. A. Wander et al. 2020)) are shown. Genes are arranged based on their pathway and include all genes with 2 or more known oncogenic mutations in the cohort. Clinical parameters shown include trial treatment information (trial treatment duration, clinical benefit and best response by RECIST 1.1, reason for discontinuation of treatment), prior CDK4/6i treatment information (CDK4/6i received, antiestrogen agent used in combination, phenotype based on prior CDK4/6i response), receptor status (biopsy-level, at primary diagnosis, and at metastatic diagnosis), timing of biopsy relative to metastatic diagnosis, and biopsy site. Research-based PAM50 subtype (when RNA-seq data is available) and tumor mutational burden of each biopsy are also shown.

For the baseline tumor biopsies (annotated as T1), WES or RNA-seq was successfully performed and passed quality-control for 18 samples each, with 14 samples having both WES and RNA-seq data (Fig. 2A). For ctDNA, WES was successfully performed and passed quality-control for 17 ctDNA samples (12 at baseline, annotated as BB1, and 5 while on therapy, annotated as BBn) from 12 patients, with 11 patients having WES from both a baseline tumor and ctDNA sample. For the pre-CDK4/6i tumor biopsies (annotated as A1), WES or RNA-seq was successfully performed and passed quality-control for 6 and 9 samples, respectively, with 5 patients having WES from both a pre-CDK4/6i and a baseline biopsy. The complete dataset consisted of data from 26 patients: germline-matched WES from 24 tumor samples (19 patients) and 17 ctDNA samples (12 patients), and RNA-seq from 27 tumor samples (22 pts) (Fig. 2A).

We first focused on the genomic landscape (WES) of the baseline tumor and blood biopsies (18 T1’s, 12 BB1’s, *n* = 19 pts with either a T1 or BB1) (Fig. 2A, Supplemental Table S7, Supplemental Table S8). We identified genomic alterations spanning the spectrum of known genes and pathways previously identified to confer resistance to CDK4/6 inhibitors in 58% (*n* = 11/19) of patients (O’Leary et al. 2018; Z. Li et al. 2018; Yang et al. 2017; S. A. Wander et al. 2020; Costa et al. 2020) and to endocrine therapy in 74% (*n* = 14/19) of patients (Razavi et al. 2018; Bertucci et al. 2019; Nayar et al. 2019; Mao et al. 2020; Hanker, Sudhan, and Arteaga 2020). These pathways, genes, and alterations include: *PTEN* bi-allelic inactivations (*n* = 1/19, 5% of pts) and *AKT1* activating mutations (*n* = 4/19, 21%) in the PI3K/AKT/mTOR pathway; *NF1* bi-allelic inactivations (1/19, 5%) in the MAPK pathway; *ERBB2* activating mutations (*n* = 2/19, 11%), *FGFR1* activating mutations (*n* = 1/19, 5%) or high amplifications (5/19, 26%), and *FGFR2* high amplifications (*n* = 1/19, 5%) in RTKs; *RB1* bi-allelic inactivations (*n* = 2/19, 11%) and *AURKA* high amplifications (*n* = 1/19, 5%) in cell cycle genes; activating mutations in *ESR1* (*n* = 9/19, 47%) in the ER pathway. In addition to these genomic alterations, we also observed loss of ER expression (as measured by IHC) in baseline tumor samples (*n* = 3/19, 16%); note that we refer to this as “ loss of ER expression” because these patients were previously diagnosed with ER+/HER2-metastatic breast cancer.

In addition to the alterations in these known resistance genes and pathways, we identified three novel potential resistance mechanisms in genes belonging to these pathways: an activating *BRAF*^V600E^ mutation (MAPK) (in patient 23, Fig. 2B), an activating *MTOR*^T1977R^ mutation (PI3K/AKT/MTOR) (in patient 22, Fig. 2C), and activating double *PIK3CA*^E545K,G1007R^ mutation (PI3K/AKT/mTOR) (in patient 19, Fig. 2D). All of these mutations were clonal in the baseline biopsy they were identified in. The activating *BRAF* mutation was not detected in the pre-CDK4/6 inhibitor biopsy, and thus was acquired/enriched following the treatment with the initial CDK4/6 inhibitor (1 year and 7 months on treatment) or the prior aromatase inhibitor (1 year and 9 months on treatment). For the other two alterations of interest (*MTOR*^T1977R^, double *PIK3CA*^E545K,G1007R^) we did not have a pre-CDK4/6i biopsy, so could not verify if they were acquired.

To further test the potential of these mutations as drivers of resistance, we looked at whether these samples have other known alterations associated with CDK4/6i resistance. For the activating *BRAF*^V600E^ mutation (patient 23), the tumor had no other clonal acquired alterations, but did show loss of ER expression, making it difficult to tease out the degree of this mutation’s resistance effect. For the activating *MTOR*^T1977R^ mutation (patient 22), the biopsy also had an *FGFR1* high amplification, which partly confounds the role of this mutation in resistance. Note that *FGFR1* amplification appears to not always drive resistance to CDK4/6i on its own, since it often co-occurs with other resistance-associated alterations (S. A. Wander et al. 2020). For the double *PIK3CA*^E545K,G1007R^ mutation (patient 19), the biopsy had an activating *ESR1* mutation, a known resistance mechanism to endocrine therapy, but did not have any known alterations associated with CDK4/6i resistance. None of these biopsies had additional known oncogenic mutations in the PI3K/AKT/mTOR, MAPK, or RTK pathways (patient 22 had an *FGFR1* high amplification, but no known oncogenic mutations), known oncogenic mutations or high-grade CNA in cell cycle genes associated with CDK4/6i resistance (*RB1, CCNE1*, or *CDK6*), or loss-of-function alterations in *FAT1*. Overall, the alterations co-occurring with the mutations we identified as potential resistance mechanisms are consistent with their proposed roles as drivers of CDK4/6i resistance, even if some of the co-occurring alterations confound their effect.

### Baseline ctDNA identified actionable genomic alterations not found in baseline tumor samples

For 11 patients, we obtained WES of both baseline tumor biopsies and baseline ctDNA. We leveraged this redundancy in our genomic data to verify the consistency between the tumor and ctDNA WES, and to see what additional information we could learn from having WES from both sources. Focusing on genes in pathways associated with resistance to endocrine therapy and CDK4/6i, we found few differences between the genomic alterations in 9 patients (*n* = 9/11, 82%) (Supplemental Fig. S1). Most of the differences we identified in these 9 patients were consistent with the higher sensitivity expected from tumor biopsies as compared to blood biopsies, particularly for CNAs. For example, the loss-of-function *RB1* mutations in patient 10 (subclonal) and 37 (clonal) were identified in the tumors but not in the ctDNA samples. In addition, we found evidence of biallelic inactivation (loss of heterozygosity and a loss-of-function mutation) for multiple tumor suppressors (*PTEN, TP53, NF1*) in the tumor but not in ctDNA samples, in which we could only identify the loss-of-function mutations. There were a few cases where some alterations were identified in the ctDNA samples but not the tumors in these 9 patients, all of which involved subclonal mutations or CNAs, and which include a subclonal *TP53* loss-of-function mutation and *FGFR1* amplifications in 3 samples. We also looked for differences between the identified genomic alterations when looking at mutations with a known oncogenic effect in cancer genes and found no additional differences between the samples of these 9 patients.

For the other 2 patients (*n* = 2/11, 18%), patients 22 and 32, we identified mutually exclusive clonal driver mutations in cancer genes between the ctDNA and the tumor biopsy pair (Fig. 2C). Each sample pair shared truncal clonal mutations, indicative of a co-existence of multiple tumor lineages in the patient’s cancer. For patient 22, we found a truncal *GATA3*^M400fs^ loss-of-function mutation with a clonal activating *MTOR*^T1977R^ mutation in the ctDNA sample and a clonal activating double *ESR1*^Y537S,L536P^ mutation in the tumor sample. For patient 32, we found a truncal *PIK3CA*^H1047L^ mutation with a clonal activating *ERBB2*^L755S^ mutation in the ctDNA sample and a clonal activating *ERBB2*^L869R^ mutation in the tumor sample. The *MTOR*^T1977R^ and *ERBB2*^L755S^ mutations found only in the blood biopsies are each clinically actionable and are associated with response to mTOR inhibitors like everolimus (Grabiner et al. 2014; Wagle et al. 2014; Kwiatkowski et al. 2016; Xu et al. 2016) and the pan-HER kinase inhibitor neratinib, respectively (Hyman et al. 2018; Bose et al. 2013). Based on this and additional evidence, OncoKB classifies *MTOR*^T1977R^ as a mutation with compelling biological evidence (OncoKB Level 4) and *ERBB2*^L755S^ as a mutation with compelling clinical evidence (OncoKB Level 3a). Thus, we identified clinically actionable clonal mutations in the ctDNA but not in the tumor baseline biopsy of 2 patients that are each likely to be the mechanism of resistance.

### Consistent transcriptomic features in genes associated with resistance to CDK4/6 inhibitors and endocrine therapy

The genomic landscape of baseline tumor biopsies in this trial spanned the spectrum of genes and pathways (RTK, MAPK, PI3K/AKT/mTOR, cell cycle, and ER pathways) known to be associated with resistance to CDK4/6 inhibitors and endocrine therapy (Fig. 2). Motivated by this finding, we hypothesized that genomic alterations in these resistance genes and pathways (in particular, known oncogenic mutations and high-grade CNAs) would have a corresponding high level of transcriptional signature activity (for oncogenic mutations and possibly for high-grade CNAs) or gene expression (for genes with high-grade CNAs). We also hypothesized that some of these genomic alterations and transcriptional signatures could correlate with the intrinsic molecular subtype (PAM50) of tumor samples. To test these hypotheses, we leveraged the genomic and transcriptomic data of the baseline tumor biopsies (14 biopsies with both WES and RNA-seq). Because of the modest size of or cohort, we focused on the genes we and others had previously identified to be associated with resistance (those in Fig. 2), transcriptional signatures from the 50 Hallmark gene sets, and 3 RTK transcriptional signatures from our recent work on resistance to endocrine therapy (Nayar et al. 2019; Mao et al. 2020). These 3 RTK signatures are associated with transcriptional activity of HER2 mutants (HER2 MUT), FGFR (FGFR ACT), or a combination of both signatures (RTK ACT).

In order to quantify whether the expression of a gene or the activity of a transcriptional signature has a high or low value, we needed a cohort to serve as a reference for gene expression. Given that these tumor samples come from patients with HR+/HER2-MBC, we also needed a reference gene expression cohort that is receptor status-balanced in order to accurately assign a molecular subtype (research-grade PAM50) (A. Prat and Parker 2020). For these purposes, we used the Metastatic Breast Cancer Project (MBCProject), which has genomic (WES, *n* = 379 tumor samples), transcriptomic (RNA-seq, *n* = 200 tumor samples) and clinical data (including receptor status: 84 HR+/HER2-, 27 HR+/HER2+, 10 HR-/HER2+, 12 HR-/HER2-), as the reference cohort (Wagle et al. 2016). After assigning a molecular subtype to the 14 biopsies in this trial, we found that 2 samples had a Normal PAM50 subtype, which is indicative of a low tumor content, so we excluded these samples from the joint genomic and transcriptomic analysis (resulting in *n* = 12 biopsies with both WES and RNA-seq and a non-Normal PAM50 subtype).

In agreement with our hypothesis, joint genomic and transcriptomic analysis revealed a high degree of consistency between the presence of known oncogenic mutations and the activity of transcriptional signatures of these pathways (Fig. 3A, Supplemental Fig. S2A). In particular, activity of each of these transcriptomic signatures was a strong classifier for the presence of oncogenic mutations in the respective pathways, as described in more detail below. Oncogenic mutations were also enriched in tumors with high activity in the transcriptomic signatures. These effects were particularly strong for *ESR1* activating mutations in the ER pathway and to a lesser degree with activating mutations in the PI3K/AKT/mTOR pathway (*AKT1* or *PIK3CA*) or the RTK/MAPK pathways (*ERBB2* or *BRAF*).

**Fig. 3.**
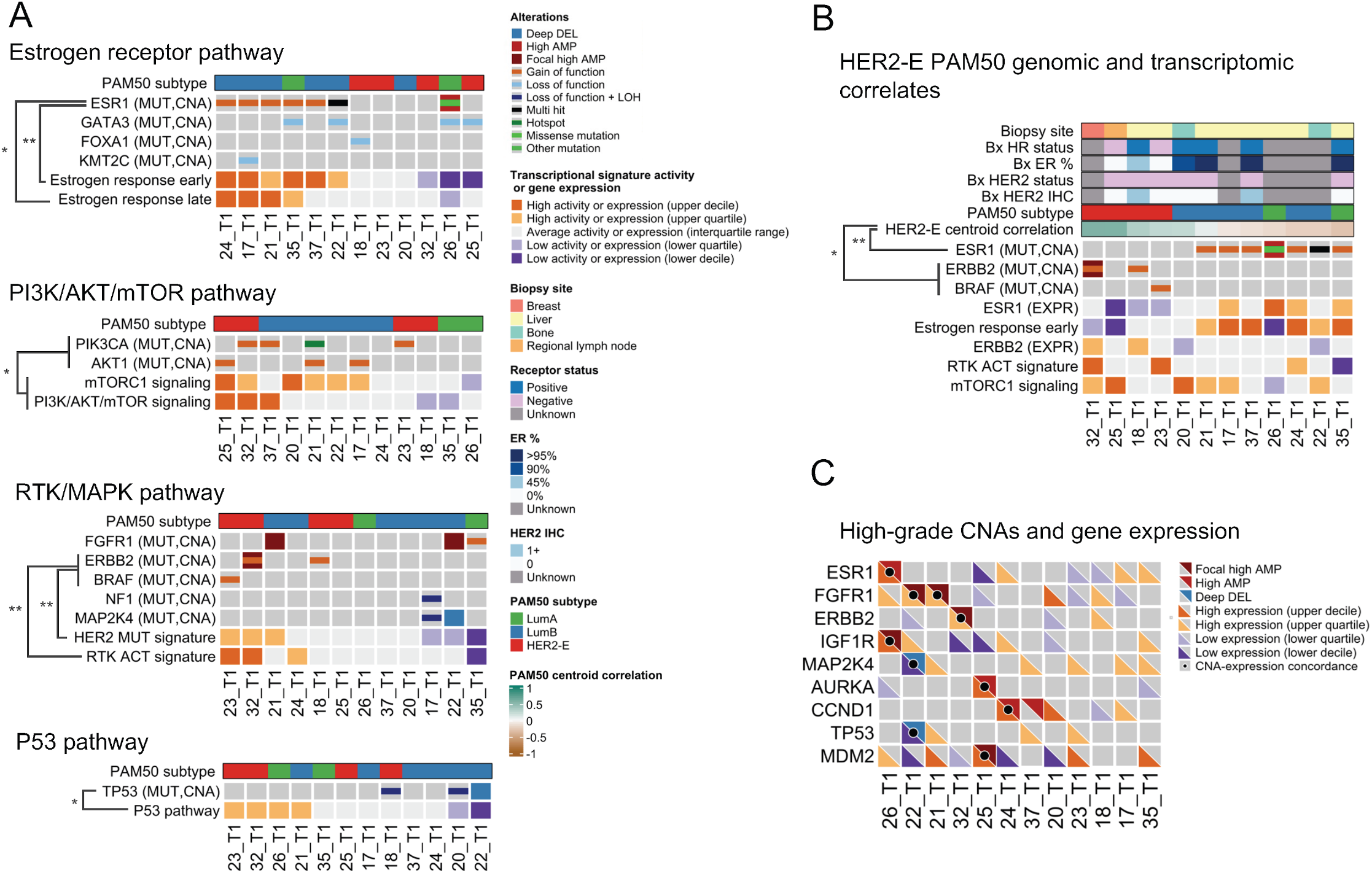
Consistency between genomic and transcriptomic features in genes associated with CDK4/6 inhibitors and antiestrogen treatment resistance. Joint genomic and transcriptomic analysis was performed on all baseline trial tumor biopsies with both WES and RNA-seq and a non-Normal PAM50 subtype (*n* = 12 samples and patients). (A) Comutation plot (CoMut) showing the consistency between the presence of oncogenic alterations and the activity of transcriptional signatures of their associated signaling pathway. Distinct genes and signatures are shown depending on the pathway (ER, PI3K/AKT/mTOR, RTK/MAPK, and P53). For each pathway, only genes from Fig. 2 with at least one known oncogenic mutation in the samples with transcriptomic data are shown. Biopsies are ordered based on the combined activity of the pathway signatures. (B) CoMut dislplaying the association between the presence of oncogenic mutations in *ERBB2* or *BRAF* and a HER2-enriched subtype, and oncogenic mutations in *ESR1* and a Luminal A or B subtype. Biopsies are ordered based on their correlation to the HER2-enriched centroid. Additional features shown are clinical and RNA-seq-based measures of ER and HER2 activity (HR and HER2 receptor status, ER percentage by IHC, HER2 IHC score, *ESR1* and *ERBB2* gene expression, and activity of the RTK ACT and estrogen response early transcriptional signatures) and biopsy site. An expanded version of panels A and B with additional clinical, genomic, and transcriptomic features is included in Supplemental Fig. S2. (C) CoMut showing the concordance between high-grade CNA and gene expression levels. Cases with CNA and gene expression concordance (high amplification or focal high amplification and upper quartile or decile expression; deep deletion and lower quartile or decile expression) are shown with a black dot. Quantiles for transcriptional signature activity and gene expression levels are derived from MBCProject. Statistically significant associations between signature activities and known oncogenic mutations are denoted with asterisks (one-sided Mann–Whitney test). (*) *P* < 0.05, (**) *P* < 0.01.

For the ER pathway, activity in the ER pathway Hallmark signatures were a strong classifier for the presence of *ESR1* activating mutations (estrogen response early, *AUC* = 1.00, *P* = 1.08 × 10^−3^; estrogen response late *AUC* = 0.86, *P* = 2.06 × 10^−2^, one-sided Mann–Whitney test) (Fig. 3A top). *ESR1* activating mutations (*n* = 6/12, 50% of biopsies) were enriched in tumors with high activity in the estrogen response early signature (*n* = 6/12, 50%; *P* = 2.16 × 10^−3^, two-sided Fisher exact test). This effect was similar but not statistically significant for the estrogen response late signature (*n* = 6/12, 50%; *P* = 6.06 × 10^−2^, two-sided Fisher exact test).

For the PI3K/AKT/mTOR pathway, combined activity of mTORC1 signaling and PI3K/AKT/mTOR signaling signatures was statistically significant as a classifier for the presence of grouped activating PI3K/AKT/mTOR pathway mutations (only *AKT1* and *PIK3CA* mutations for these samples) (*AUC* = 0.83, *P* = 3.25 × 10^−2^, one-sided Mann–Whitney test) (Fig. 3A middle top). None of these comparisons were statistically significant when looking at each of these signatures or activating mutations individually, or when looking at tumors with high activity in these signatures (all comparisons *P* > 5.00 × 10^−2^), although activity in the mTORC1 signaling signature was borderline statistically significant as a classifier for activating *AKT1* mutations (*AUC* = 0.85, *P* = 5.00 × 10^−2^, one-sided Mann–Whitney test).

For the RTK/MAPK pathway, activity of the RTK ACT or HER2 MUT signatures was each a good classifier for the presence of grouped activating *ERBB2* or *BRAF* mutations (*AUC* = 0.96, *P* = 9.09 × 10^−3^, one-sided Mann–Whitney test for each signature) (Fig. 3A middle bottom). None of these comparisons were statistically significant when taking each of these activating mutations individually and there was no statistically significant enrichment of these mutations when looking at tumors with high activity in RTK/MAPK signatures (all comparisons *P* > 5.00 × 10^−2^). Other known RTK/MAPK alterations (*NF1* biallelic inactivation, *FGFR1* activating mutations and high amplifications) were not associated with high activity in the RTK/MAPK signatures and seemed to inversely correlated with these signatures (Fig. 3A, Supplemental Fig. S2). As detailed below, the lack of association with RTK/MAPK signatures in these individual cases could be related to the presence of concurrent activating *ESR1* mutations in those tumors (17_T1, 21_T1, 22_T1, 35_T1).

As an additional consistency check, we verified that deactivating *TP53* alterations (biallelic inactivation, *n* = 2/12, 17%; deep deletions, *n* = 1/12, 8%) were associated with P53 pathway activity (Fig. 3A bottom). Activity in the P53 pathway signature was a statistically significant classifier for the presence of grouped *TP53* biallelic inactivation and deep deletions (*AUC* = 0.93, *P* = 1.81 × 10^−2^, one-sided Mann–Whitney test). Similarly, these *TP53* alterations were enriched in samples with low P53 pathway signature activity (*P* = 4.54 × 10^−2^, two-sided Fisher exact test).

Given the observed association of activating *ERBB2* and *BRAF* mutations with RTK/MAPK pathway transcriptional signature, we hypothesized this association could be related to the PAM50 subtype of these tumor samples. Consistent with this hypothesis, grouped *ERBB2* and *BRAF* mutations were exclusive to tumors with a HER2-Enriched (HER2-E) PAM50 subtype (*n* = 4/12, 33%), in which they were enriched (*P* = 1.82 × 10^−2^, two-sided Fisher exact test), and correlation to the HER2-E PAM50 centroid was a good classifier for the presence of these mutations (*AUC* = 0.93, *P* = 1.82 × 10^−2^, one-sided Mann–Whitney test) (Fig. 3B). In addition, correlation to the HER2-E PAM50 centroid was a good classifier for the absence of activating *ESR1* mutations (*AUC* = 0.92, *P* = 7.58 × 10^−3^, one-sided Mann–Whitney test). Unlike the HER2-E PAM50 centroid, the RTK ACT or HER2 MUT signatures were not statistically significant classifiers for activating *ESR1* mutations (*P* > 5.00 × 10^−2^). Activating *ESR1* mutations were exclusive to samples with a Luminal A (*n* = 2/12, 17%) or B (*n* = 6/12, 50%) subtype and were absent in the tumors with *ERBB2* and *BRAF* mutations. Mutual exclusivity between activating mutations in *ESR1* and *ERBB2* or *BRAF* is consistent with prior work in HR+/HER2-MBC (Razavi et al. 2018; Nayar et al. 2019; Mao et al. 2020; S. A. Wander et al. 2020). To our knowledge, this is the first report that the presence of activating mutations in the RTK/MAPK pathway or *ESR1* has been associated with the PAM50 molecular subtype of a tumor in HR+/HER2-MBC.

To verify the consistency between high-grade CNAs (high amplifications, deep deletions) and gene expression, we looked at whether tumors with high-grade CNAs had a corresponding high or low level of gene expression. For 9 out 10 high-grade CNAs there was a corresponding high or low level of expression in these genes (Fig. 3C), which included *FGFR1* (*n* = 2/12, 17%), *ESR*1 (*n* = 1/12, 8%), *ERBB2* (*n* = 1/12, 8%), *IGF1R* (*n* = 1/12, 8%), and *AURKA* (*n* = 1/12, 8%), among others. Conversely, for the majority of cases with an above-average level of expression in these genes there were no corresponding high-grade CNAs (52 total gene/tumor cases, 9 of which had a high-grade CNA), a result that is consistent with recent work (Sánchez-Guixé et al. 2022). These results support the often-used assumption that high-grade CNAs in resistance-associated genes have a strong effect on their expression but highlight how above-average expression levels do not often have an associated high-grade CNA.

In summary, we found that multiple genomic resistance mechanisms in the ER, RTK/MAPK, and PI3K/AKT/mTOR pathways, such as activating mutations in *ESR1, ERBB2*, or *AKT1* and *FGFR1* amplifications, have specific transcriptomic features (pathway transcriptional signatures, gene expression levels) associated with them. Specifically, there was a statistically significant association between *ESR1* mutations and ER pathway Hallmark signatures, *ERBB2*/*BRAF* mutations and the RTK ACT or HER2 MUT signatures, and the presence of *ERBB2*/*BRAF* or absence of ESR1 mutations and the HER2-E centroid correlation. These results suggest that these transcriptomic features could be used to identify the pathways driving CDK4/6i and endocrine resistance in a tumor.

### Combined genomic and transcriptomic features in baseline biopsies identify likely mechanism of resistance to prior CDK4/6 inhibitor and endocrine therapy treatments in nearly every patient

The genomic and transcriptomic analysis of baseline biopsies identified known and new potential features associated with the resistance to CDK4/6 inhibitors and endocrine therapy. These features include activating mutations in *ESR1, ERBB2, FGFR1, AKT1, BRAF*, and *MTOR*; double *PIK3CA* activating mutations; loss-of-functions mutations in *NF1, RB1*, and *PTEN*; amplifications in *FGFR1, FGFR2, ERBB2*, and *AURKA*; ER loss; and transcriptional features such as the HER2-E subtype, high transcriptional activity ER signatures, or high RTK activity (high expression of RTKs or high RTK signature activity).

Based on these and additional features, we asked whether these known and plausible resistance mechanisms could explain each baseline tumor’s resistance to prior CDK4/6i and antiestrogen treatments. Additional features we considered include known resistance features such as the Basal PAM50 subtype (Aleix Prat et al. 2021), and plausible resistance features, some of which have preliminary studies that associate them to reduced sensitivity to CDK4/6i and endocrine therapy, such as amplification or high expression of *IGF1R* or *INSR* (Seth A. Wander, Mao, et al. 2021; Fox et al. 2011; Mao et al. 2020), and low ER activity (low *ESR1* expression or low ER pathway signature activity) (O’Leary et al. 2018).

For 22 out of 23 patients with WES and/or RNA-seq of a baseline biopsy (*n* = 22/23, 96%), we identified genomic or transcriptomic features that could explain the tumor’s resistance to CDK4/6i or anti-ER treatments (Fig. 4, Supplemental Fig. S3, Supplemental Table S9). For 14 patients (*n* = 14/23, 61%) there were known resistance mechanisms to both of these treatments (i.e., published evidence of a mechanistic role in resistance to these treatments), for 3 patients (*n* = 3/23, 13%) there was a combination of known and plausible mechanisms, and for 5 patients (*n* = 5/23, 22%) there were only plausible mechanisms (i.e., preclinical or theoretical evidence suggesting a role in resistance). Given that the resistance mechanisms identified for each patient can be based on clinical, genomic, or transcriptomic features, we looked at how often each explained the tumor’s treatment resistance. In particular, we focused on the cases where oncogenic mutations in resistance genes, clinical features, or transcriptomic features alone could explain treatment resistance.

**Fig. 4.**
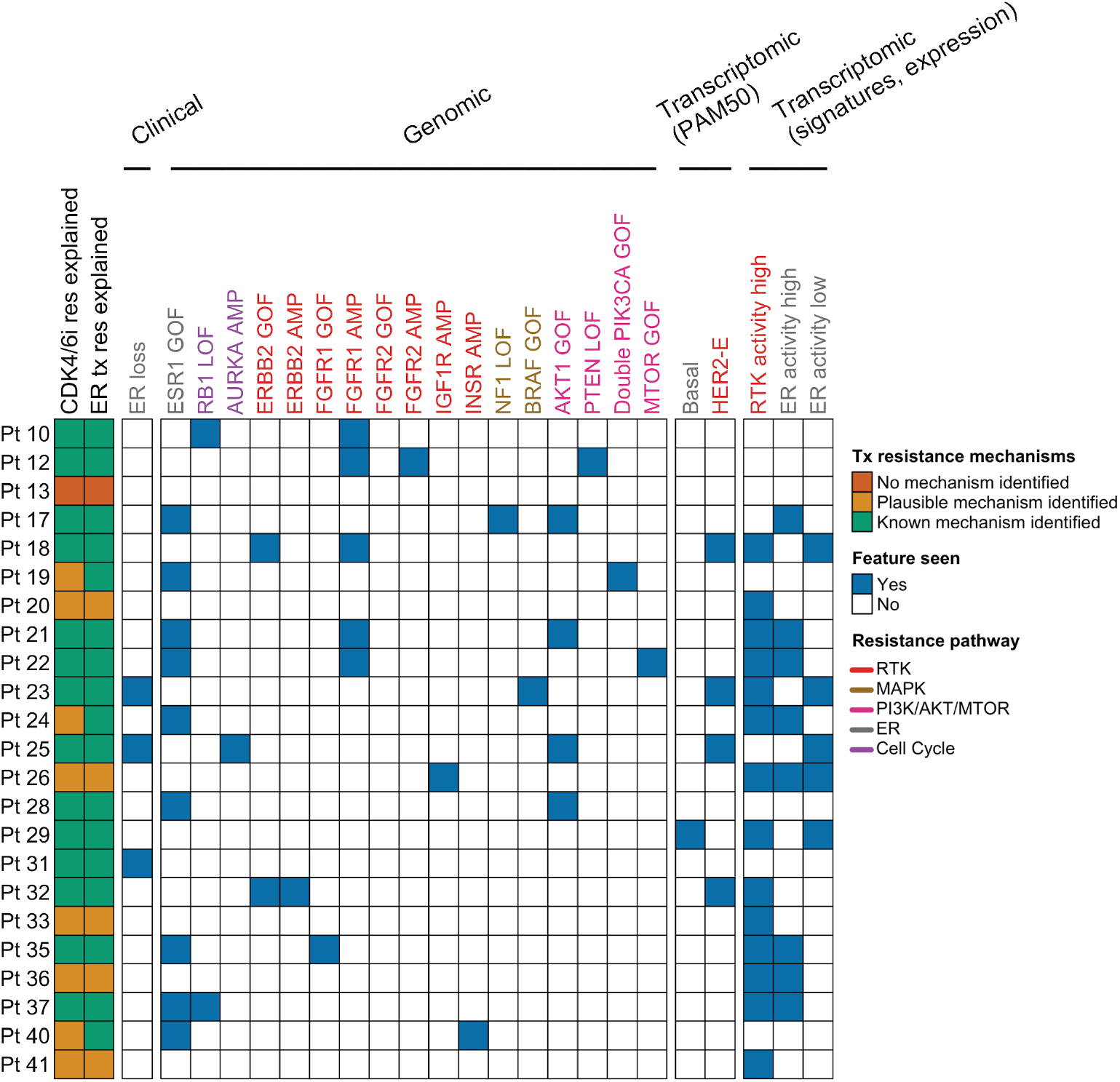
Clinical, genomic, and transcriptomic features can explain resistance to CDK4/6 inhibitors and antiestrogen treatment in patient’s tumors. The chart encodes whether key features associated with resistance to CDK4/6 inhibitors or antiestrogen treatment are seen in the trial baseline biopsies of patients with either WES or RNA-seq of a baseline sample (*n* = 23 patients). For each patient, it includes whether these features could explain if the baseline tumor is resistant to prior CDK4/6i and antiestrogen treatments, and if these are known or plausible resistance mechanisms. For 22 out of 23 patients a known or plausible resistance mechanism was identified. Features included are based on clinical assays (loss of ER measured by IHC), genomic data (known oncogenic mutations and high-grade amplifications in known or plausible resistance genes), and transcriptomic data (Basal subtype, HER2-enriched subtype, high expression or activity in an RTK gene or RTK signature, high/low expression or activity in *ESR1* or an ER pathway signature). Features are colored based on the resistance signaling pathway they are associated with. A modified version of this figure that groups these features by gene and signaling pathway, and includes the data types available for each patient is included in Supplemental Fig. S3. GOF, gain of function mutation; LOF, loss of function mutation; AMP, high amplification.

For 13 patients (*n* = 13/23, 57%), resistance to both treatments could be explained solely by considering oncogenic mutations in known (*ESR1, AKT1, PTEN, NF1, FGFR1, ERBB2, RB1*) or plausible resistance genes (*BRAF, MTOR*, double mutations in *PIK3CA*), or ER loss (pts. 23, 25, and 31). For 5 patients (*n* = 5/23, 22%; 4 with no genomic data), the resistance mechanisms identified were solely transcriptomic features: Basal PAM50 subtype (along with high RTK activity and low ER activity) in pt. 29, high RTK activity (with high *IGF1R* expression) and ER activity (high estrogen response early signature activity) in pt. 36, and high RTK activity in pts. 20 (with high *FGFR1*), 33 (with high *IGF1R* expression), and 41 (with high activity of the RTK ACT and HER2 MUT signatures). For 4 patients (*n* = 4/23, 17%), the resistance mechanisms identified included a combination of oncogenic mutations, high-grade CNAs, and transcriptomic features. For example, *FGFR1* amplification and *RB1* loss-of-function for pt. 10; an activating *ESR1* mutation and high activity of the RTK ACT signature for pt. 24; and *IGF1R* amplification and high *IGF1R* expression, a *GATA3* loss-of-function mutation, and low ER activity (low signature activity of the estrogen response early and late signatures) for pt. 26.

In summary, we found that in 22 out of 23 patients we could identify known or plausible resistance mechanisms to the prior CDK4/6i and antiestrogen treatments by using the clinical, genomic, and transcriptomic features in their baseline tumors. This includes 5 patients in which the only mechanism identified was a transcriptomic feature, illustrating how transcriptomic data can provide complementary information not present in genomic data and provide insights even for cases when no genomic data is available.

### Evolutionary analysis revealed convergent and divergent paths to CDK4/6i + anti-ER treatment resistance in tumors with distinct lineages

For 6 patients for whom we had WES from paired pre-treatment and post-resistance tumor samples (5 patients) or concurrent post-treatment tumor and ctDNA samples from distinct tumor lineages (2 patients), we wanted to identify the genomic alterations that were acquired or became enriched following the prior CDK4/6i treatment and that could be driving treatment resistance. To do this, we carried out evolutionary analysis (tumor phylogeny, clonal dynamics) to identify acquired or enriched genomic alterations in the trial baseline sample(s) (which are post-CDK4/6i) as compared to the older, pre-CDK4/6i sample (Figs. 5 and 6). To help contextualize the treatments these samples have been exposed to (and, putatively, become resistant to), we also included a clinical case history for these patients (treatment sequence and duration, timing of biopsies and diagnoses) (Supplemental Table S10).

**Fig. 5.**
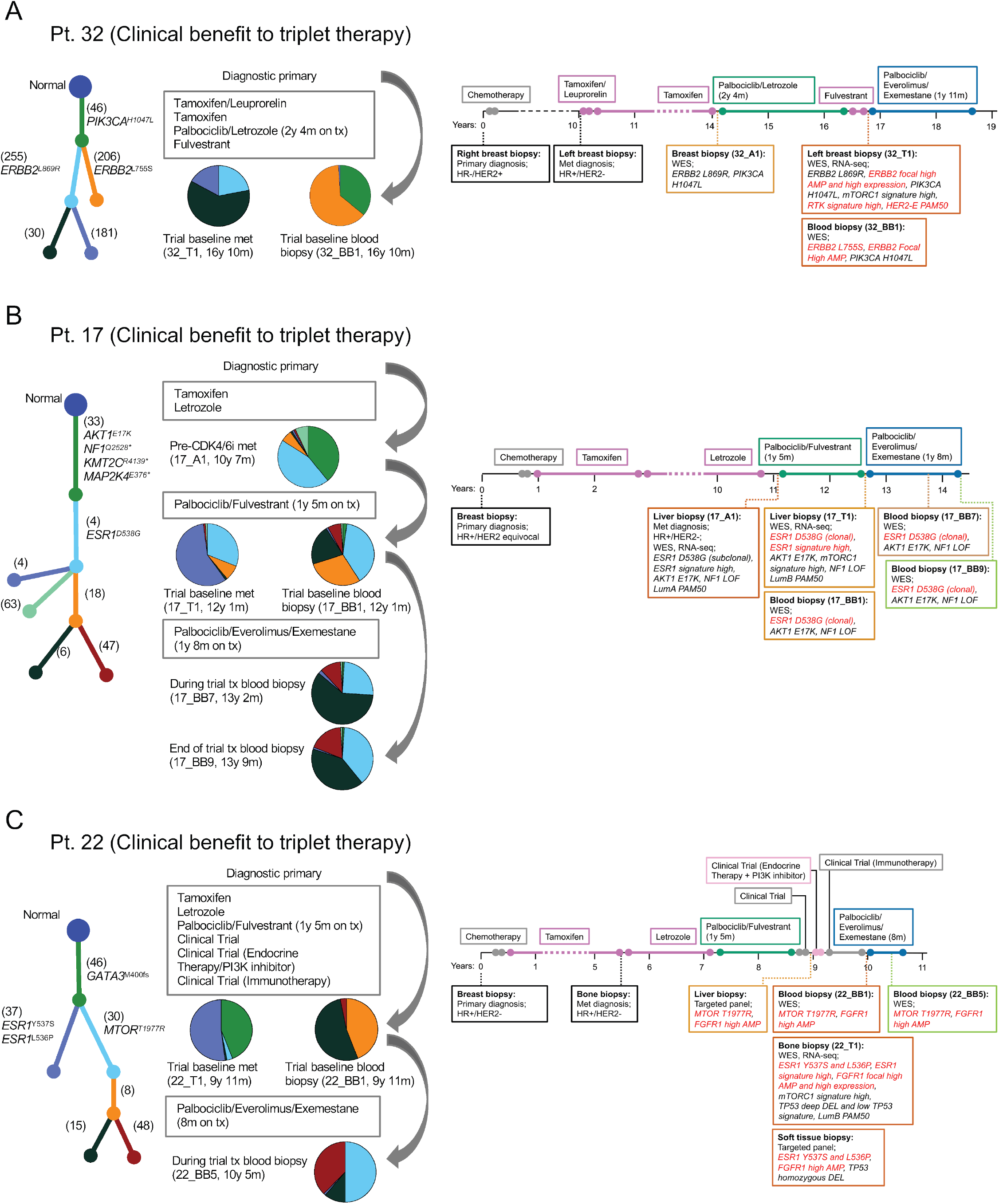
Tumor evolutionary analysis and clinical vignettes for patients who derived clinical benefit from palbociclib, everolimus, and exemestane triplet therapy. Analysis of tumor phylogeny and clonal dynamics following CDK4/6i and anti-ER therapy revealed convergent (HER2 activation) and divergent (ER or PI3K/AKT/mTOR activation) paths to treatment resistance in tumors with distinct lineages. (A, B) Evolutionary analysis, acquired genomic alterations to prior CDK4/6i, and treatment history for patients with pre-CDK4/6i and post-CDK4/6i (trial baseline) biopsies. Convergent evolution of *ERBB2* activation in two distinct co-existing tumor lineages is shown in panel A (an activating clonal *ERBB2*^L869R^ mutation and an acquired *ERBB2* focal high amplification in 32_T1; an acquired activating clonal *ERBB2*^L755S^ mutation in 32_BB1). Increase in clonality of an activating *ESR1*^D538G^ mutation is shown in panel B (from subclonal to clonal). (C) A case with two distinct co-existing tumor lineages in its post-CDK4/6i biopsies, each with clonal drivers mutations in divergent pathways (ER pathway with an activating clonal *ESR1*^Y537S,L536P^ mutation in 22_T1; PI3K/AKT/mTOR pathway with an activating clonal *MTOR*^T1977R^ mutation in 22_BB1). Even though no pre-CDK4/6i biopsy was available for this patient, targeted panel data from other post-CDK4/6i biopsies confirmed the existence of these tumor lineages. In a patient’s tumor phylogenic tree, each subclone is associated with a branch and a color, and this color matches that in the pie chart that quantifies the relative abundance of each subclone in the tumor. The number of mutations unique to each subclone and known oncogenic mutations are shown next to each branch. Data shown in the clinical vignettes includes the timing of treatments and biopsies, and selected clinical, genomic, and transcriptomic features. Time on treatment for CDK4/6i-containing therapies are included in the figure. Acquired genomic alterations (in A and B) or putatively acquired genomic alterations (in C) following CDK4/6i therapy are shown in red, together with their associated transcriptomic features.

**Fig. 6.**
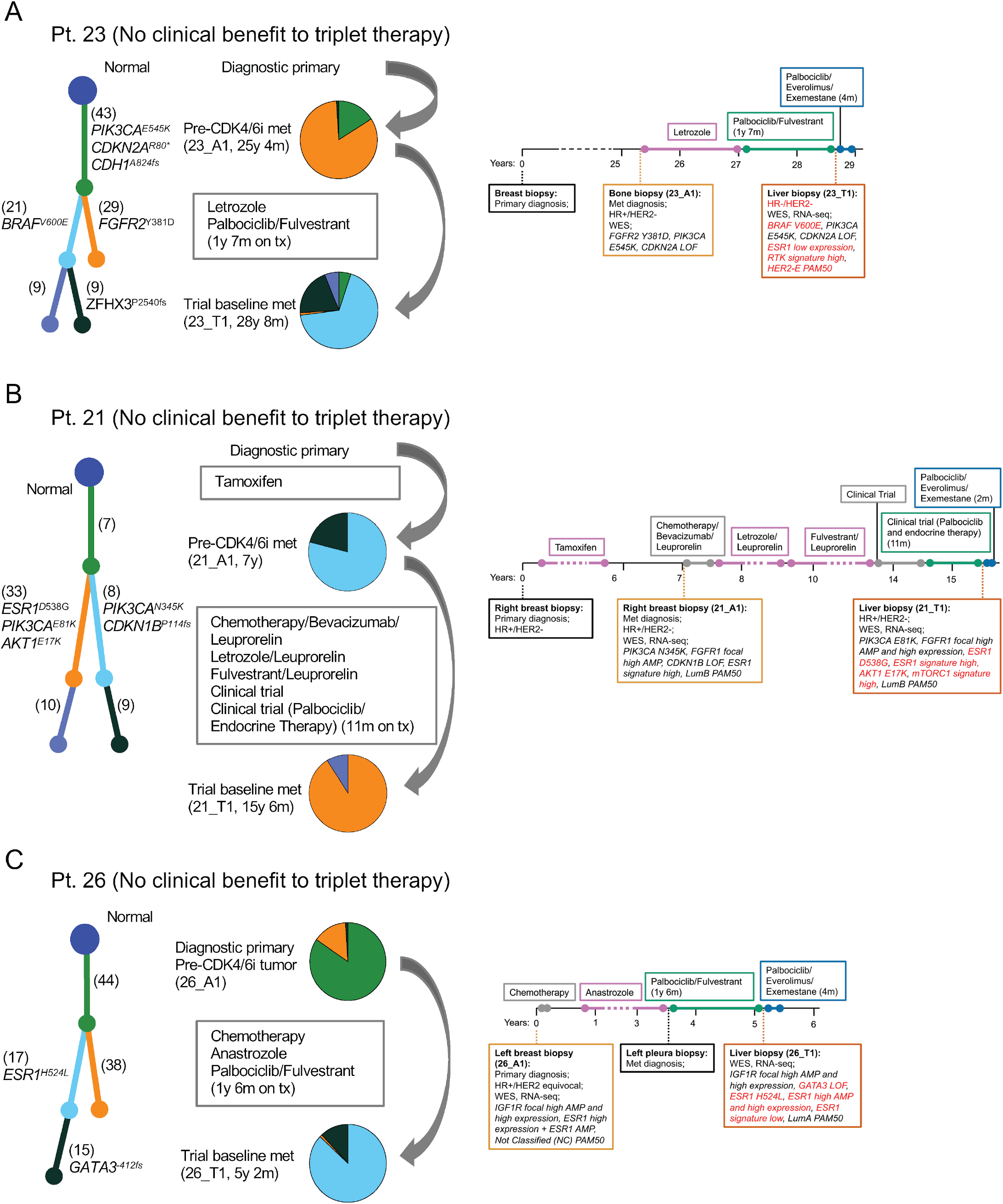
Tumor evolutionary analysis and clinical vignettes for patients who did not derive clinical benefit from palbociclib, everolimus, and exemestane triplet therapy. Evolutionary analysis, acquired genomic alterations to prior CDK4/6i, and treatment history are shown for these patients. (A) Acquired clonal activating *BRAF*^V600E^ mutation concurrent with ER loss. Transcriptomic features (low *ESR1* expression, HER2-enriched PAM50, high RTK signature activity) are concordant with these acquired events. (B) Acquired clonal activating *AKT*^E17K^ and *ESR1*^D538G^ mutations. (C) Acquired clonal *ESR1*^H524L^ mutation (variant of unknown significance), an acquired subclonal *GATA3*^*-412fs*^ truncating mutation, and an increased *ESR1* amplification (from amplification to high amplification). In a patient’s tumor phylogenic tree, each subclone is associated with a branch and a color, and this color matches that in the pie chart that quantifies the relative abundance of subclones. The number of mutations unique to each subclone and known oncogenic mutations are shown next to each branch. Data shown in the clinical vignettes includes the timing of treatments and biopsies, and selected clinical, genomic, and transcriptomic features. Time on treatment for CDK4/6i-containing therapies are included in the figure. Acquired genomic alterations following CDK4/6i therapy are shown in red, together with their associated transcriptomic features.

For all patients with a pre-CDK4/6i sample, we identified acquired or enriched genomic alterations in known resistance genes. For patient 32, the post-resistance tumor sample had a clonal *ERBB2*^L869R^ activating mutation that was also present in the pre-treatment sample, as well as an acquired *ERBB2* focal high amplification (copy number above ploidy from 1.6 to 9.7) not found in the pre-treatment sample. We did not detect *ERBB2*^L869R^ in the concurrent post-resistance ctDNA sample, and instead identified an acquired clonal *ERBB2*^L755S^ activating mutation that was not found in the pre-treatment sample (Fig. 5A). For patient 17, the post-resistance sample showed an enrichment in an activating *ESR1*^D538G^ mutation, which was subclonal in the pre-CDK4/6i samples but clonal in the post-CDK4/6i sample (Fig. 5B). For patient 23, the post-resistance sample had shared truncal mutations (*PIK3CA*^*E545K*^, *CDH1*^*A824fs*^, and others) and an acquired clonal *BRAF*^V600E^ activating mutation (Fig. 6A). For patient 21, the post-resistance sample had shared truncal alterations (mutations of unknown significance and a *FGFR1* focal high amplification) and an acquired clonal *ESR1*^D538G^ and *AKT1*^*E17K*^ activating mutations (Fig. 6B). For patient 26, the post-resistance sample had shared truncal alterations (SNVs of unknown significance and an *IGF1R* focal high amplification), an acquired subclonal *GATA3*^*-412fs*^ truncating mutation, an acquired clonal *ESR1*^*H524L*^ missense mutation, and an increased amplification in *ESR1* (copy number above ploidy from 5.9 to 7.6) that included the region with *ESR1*^*H524L*^ (Fig. 6C).

Of these patients, two had acquired or enriched genomic alterations only in the ER pathway (*ESR1*^D538G^ in pt. 17, Fig. 5B; *GATA3*^*-412fs*^, *ESR1*^*H524L*^, and *ESR1* high amplification in pt. 26, Fig 5C) and no known acquired alterations associated with CDK4/6i resistance, which suggests that the mechanisms of resistance is primarily related to the endocrine partner.

For patients 22 and 32, for which we had identified mutually exclusive clonal driver mutations in their baseline blood and tumor biopsies (Fig. 2C), evolutionary analysis confirmed the co-existence of multiple tumor lineages in each of these cancers (Fig. 5A, 5C).

For patient 32, the identified tumor lineages showed convergent evolution of *ERBB2* activation through distinct known activating mutations and the focal high amplification of *ERBB2* (Fig. 5A). The baseline tumor sample is dominated by a lineage with *ERBB2*^L869R^ and the baseline blood sample by a lineage with *ERBB2*^L755S^, both sharing a truncal *PIK3CA*^H1047L^ mutation. The pre-CDK4/6i sample had clonal *ERBB2*^L869R^ and *PIK3CA*^H1047L^ mutations, and thus, shared the same lineage as the baseline tumor sample. Unlike the baseline samples, the pre-CDK4/6i tumor did not show an amplification of *ERBB2*, consistent with the original HR+/HER2-MBC diagnosis. The activation of *ERBB2* in the baseline tumor is also reflected in its transcriptional features: a HER2-E PAM50 subtype and high activity in the RTK ACT and HER2 MUT signatures. To our knowledge, this is the first reported case of convergent evolution of *ERBB2* activation following CDK4/6i and endocrine therapy treatment.

For patient 22, we identified two tumor lineages with clonal drivers mutations in divergent pathways: a double *ESR1*^Y537S,L536P^ mutation (ER pathway) in the tumor sample and an *MTOR*^T1977R^ mutation (PI3K/AKT/mTOR pathway) in the ctDNA sample (Fig. 5C). Both samples had a shared truncal *GATA3*^M400fs^ mutation and a shared *FGFR1* high amplification, which was additionally classified as a focal high amplification for the tumor sample. Each of the baseline samples had known or plausible resistance mechanisms to CDK4/6 inhibitors and endocrine therapy (*ESR1*^Y537S,L536P^ and *FGFR1* amplification for 22_T1; *MTOR*^T1977R^ and *FGFR1* amplification for 22_BB1). Although we could not obtain a pre-CDK4/6i sample for this patient, we did verify that both lineages were observed in metastatic biopsies taken after CDK4/6i treatment and before beginning the trial regimen. These biopsies were profiled using targeted DNA sequencing (OncoPanel) and were from distinct metastatic sites than the baseline biopsy (liver and soft tissue vs bone for the baseline biopsy). The soft tissue biopsy had a double *ESR1*^Y537S,L536P^ mutation, an *FGFR1* high amplification, and a *TP53* homozygous deletion, consistent with what we identified in the baseline tumor biopsy. The liver biopsy had an *MTOR*^T1977R^ mutation and an *FGFR1* high amplification, consistent with the ctDNA sample.

Overall, tumor evolutionary analysis identified acquired genomic alterations that we attributed to acquired CDK4/6i and endocrine therapy resistance. These acquired genomic alterations were in the RTK (*ERBB2* amplification, *ERBB2*^L755S^), MAPK (*BRAF*^V600E^), PI3K/AKT/mTOR (*AKT1*^*E17K*^), or ER (*ESR1*^*H524L*^, *ESR1*^D538G^, *GATA3*^*-412fs*^) pathways. Finally, we identified co-existing tumor lineages with distinct driver mutations in resistance genes in two patients. In one of these patients, there was strong evidence for the convergent evolution of *ERBB2* activation following CDK4/6i therapy, possibly the first reported case of this kind.

### Response to combined palbociclib, everolimus, and exemestane was correlated with activation of the mTOR pathway

We looked for genomic and transcriptomic features in the baseline biopsies (*n = 23* patients with either WES or RNA-seq of a baseline biopsy) that correlated with subsequent clinical benefit to the clinical trial of combined palbociclib + everolimus + exemestane (*n* = 4/23, 17%).

Although there were no definite genomic correlates associated with the group that derived clinical benefit, (which was not surprising given the limited sample size), we did identify alterations in the PI3K/AKT/mTOR pathway as a plausible correlate. At the gene level there were no clear group differences and many of the known mutations in resistance genes were found in both groups, including activating mutations in *ESR1, AKT1*, and *ERBB2*, and amplifications in *FGFR1* and *ERBB2* (Fig. 2). When looking at genomic alterations in pathways, we found that the tumors in the clinical benefit group all had a known oncogenic mutation in the PI3K/AKT/mTOR pathway (Fig. 7A). For patient 22, the ctDNA sample (22_BB1) had a clonal activating *MTOR*^T1977R^ mutation, which is consistent with the evidence linking this mutation with response to mTOR inhibitors like everolimus (Grabiner et al. 2014; Wagle et al. 2014; Kwiatkowski et al. 2016; Xu et al. 2016) (Figs. 2B, 5C). For patient 19, the ctDNA sample had a clonal activating double *PIK3CA*^E545K,G1007R^ mutation (19_BB1, Fig. 2D). Patients 17 and 32 had clonal activating *AKT1*^*E17K*^ and *PIK3CA*^*H1047L*^ mutations, respectively (17_T1 and 32_T1, Fig. 2B). Given that everolimus targets the PI3K/AKT/mTOR pathway, this suggests the observed association between response and PI3K/AKT/mTOR mutations could be attributed to the everolimus component of combination therapy.

**Fig. 7.**
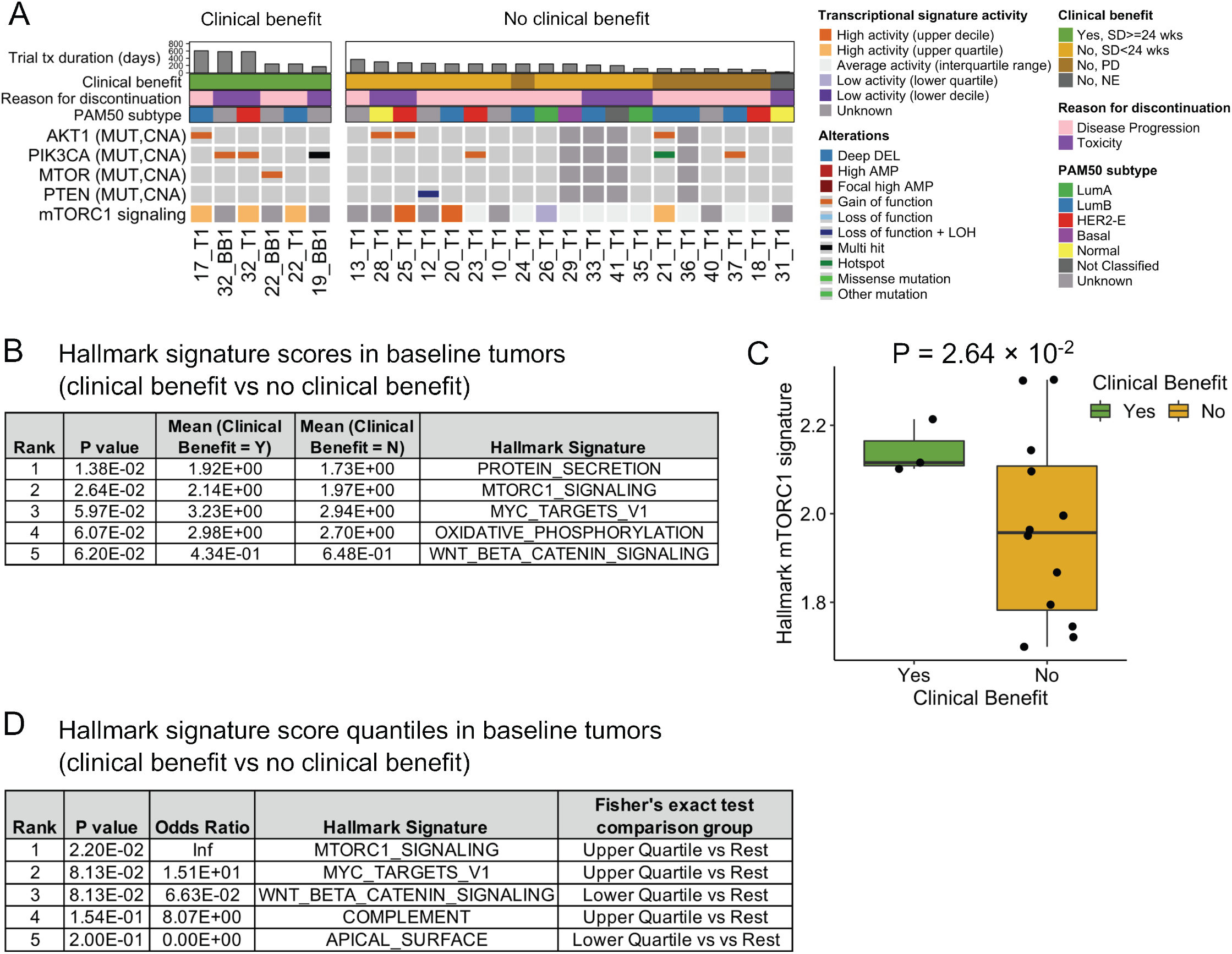
Correlation between mTOR pathway activity and response to triplet therapy in baseline tumor samples. (A) Comutation plot (CoMut) displaying the putative association between clinical benefit to triplet therapy and the presence of genomic or transcriptomic features associated with PI3K/AKT/mTOR pathway activation. Each tumor and blood biopsy sample from patients that derived clinical benefit had either a known oncogenic mutation in *AKT1, PIK3CA*, or *MTOR*, or high activity of the mTORC1 signaling signature. Of note, two out of four patients with *AKT1*^*E17K*^ mutations discontinued treatment because of toxicity. PI3K/AKT/mTOR pathway genes with at least one known oncogenic mutation are shown. Baseline tumor samples with either WES or RNA-seq are shown. Baseline blood biopsies were included when they were from a distinct lineage than the tumor biopsy (22_BB1, 32_BB2) or when there were no sequenced tumor biopsies (19_BB1). Biopsies are ordered by treatment duration on triplet therapy. (B-D) Top results from comparing Hallmark signature activity in baseline tumor biopsies with or without clinical benefit. The mTORC1 signaling signature is one of the top signatures associated with clinical benefit. Welch’s t-test (two-sided) results and Hallmark signature activity for mTORC1 signaling are shown in panels B and C, respectively. Fisher exact test (two-sided) results comparing enrichment of tumors with a Hallmark signature activity in the upper or lower quartiles are shown in panel D. The Hallmark signatures contain 50 gene sets in total. Quantiles for transcriptional signature activity are derived from MBCProject.

Given the observed association between PI3K/AKT/mTOR mutations and clinical benefit, we looked at possible reasons why this association was not observed for all the activating *AKT1*^*E17K*^ mutations. Of the four patients with an *AKT1*^*E17K*^-mutant tumor, two of them had stable disease but discontinued the trial due to toxicity (Fig. 7A). Of the other two patients, one derived clinical benefit (patient 17, stable disease > 24 weeks) while the other one did not (patient 21, best response was progressive disease). Their main noteworthy genomic/transcriptomic difference is in their HER2 MUT signature activity (high for patient 21 and low for patient 17; both were *ESR1*-mutant, RTK/MAPK mutant, and had a Luminal B subtype). Thus, we were only able to fully evaluate whether *AKT1*^*E17K*^-mutant tumors responded to combined palbociclib, everolimus, and exemestane for two cases and found a clinical benefit for one of these cases.

Consistent with the association between PI3K/AKT/mTOR mutations and clinical benefit, transcriptomic analysis identified a correlation between mTORC1 pathway activity and clinical benefit. mTORC1 signaling signature activity was higher in samples with clinical benefit (*P* = 2.64 × 10^−2^, two-sided Welch’s t-test) and was top 2 among all Hallmark signatures (Fig. 7B, 7C, Supplemental Table S11). A similar result was found when looking at enrichment of high or low activity of Hallmark signatures and clinical benefit, that is, samples with high activity in mTORC1 signaling were enriched in those associated with clinical benefit (*P* = 2.20 × 10^−2^, two-sided Fisher exact test) (Fig. 7A, 7D, Supplemental Table S11).

In summary, we found early evidence that clinical benefit to palbociclib + everolimus + exemestane is correlated with activation of the mTOR pathway, namely, PI3K/AKT/mTOR mutations and mTORC1 pathway activity.

## Discussion

In this study, we analyzed multi-omic and clinical data from patients diagnosed with metastatic HR+/HER2-breast cancer who had progressed on a prior CDK4/6 inhibitor, and were treated with a CDK4/6i-containing triplet therapy (palbociclib + everolimus + exemestane) as part of a phase 1b/2 clinical trial. This is one of the first clinical trials evaluating the benefit of continued CDK4/6 therapy after progression on a prior CDK4/6 inhibitor and, to our knowledge, is the first to carry out in-depth multi-omic tumor analysis in baseline metastatic tissue and blood samples taken after development of CDK4/6i resistance. Although the CBR of 18.8% did not meet the pre-specified threshold (CBR ≥ 65%), the multi-omic data from this trial provided a unique opportunity to study both the landscape of resistance to CDK4/6 inhibitors and correlates of response to subsequent therapy with combined palbociclib, everolimus, and exemestane. Unlike most prior studies on CDK4/6 inhibitor resistance, which focused exclusively on tumor DNA, our study explored both the DNA landscape (via WES) and the transcriptomic landscape (via RNA-seq), using both tumor biopsies and ctDNA.

### Differences in clinical benefit rate between triple therapy clinical trials and potential factors behind them

Our clinical trial results stand in contrast to the phase I/II TRINITI-1 trial, in which the triplet of ribociclib + everolimus + exemestane resulted in a CBR of 65.2% or 59.4% by arm in their final efficacy analysis (Hurvitz et al. 2022). Although both trials had relatively similar eligibility criteria, the patient population differed in important aspects. In particular, the patient population in our trial was more heavily pretreated both in terms of prior endocrine therapies (53% with 2 or more lines vs 22% in TRINITI-1) and prior chemotherapy (38% vs 13% in TRINITI-1) (Bardia et al. 2021). Concordantly, we also found a larger proportion of *ESR1* mutations at baseline in our trial (47% of patients with a known activating mutation vs 34% in TRINITI-1 with any *ESR1* mutation) (Bardia et al. 2021).

Another important difference between these trials is that the pattern of CDK4/6i re-exposure is different: in both trials a great majority of patients had previously received palbociclib (94% here vs 100% in TRINITI-1) and very few had previously received either abemaciclib or ribociclib (6% here vs 13% in TRINITI-1) (Bardia et al. 2021). This means that in our trial most patients were re-exposed to the same CDK4/6i, while in TRINITI-1 they were predominantly exposed (i.e. switched) to a different one, a notable difference that could be playing an important role in the different trial results.

Recent work on CDK4/6i re-exposure is consistent with the switch of CDK4/6i playing an important role in response to therapy. A multicenter retrospective analysis of abemaciclib exposure after progression on palbociclib found an unexpected benefit from abemaciclib, with a median PFS of 5.3 months and a treatment duration of ≥6 months for 37% of patients (Seth A. Wander, Han, et al. 2021). In MAINTAIN, a phase II randomized trial on CDK4/6i-resistant patients evaluating the switch of anti-estrogen therapy and either ribociclib or placebo, most patients also switched to a different CDK4/6i (11% of the full cohort had prior ribociclib), and they found a statistically significant improvement in the ribociclib arm compared to the placebo arm (median PFS of 5.33 vs 2.76 months, respectively) (Kalinsky et al. 2022). Although *in vitro* and *in vivo* work suggest that ribociclib and palbociclib have similar pharmacological profiles and only abemaciclib differs from them (because of its CDK2 and CDK9 activity) (Freeman-Cook et al. 2021; Hafner et al. 2019), the results of MAINTAIN, and the large difference in CBR between our trial and TRINITI-1 (18.8% vs 65.2% or 59.4%) points to the possibility that switching between distinct CDK4/6 inhibitors might be a strong contributor to this difference.

### Oncogenic signaling pathways associated to CDK4/6i resistance were recapitulated by joint genomic and transcriptomic analysis

The DNA alterations found in the trial baseline biopsies (post-CDK4/6i, pre-triplet therapy; Fig. 2) captured the spectrum of genes and pathways comprising the known genomic landscape of CDK4/6i resistance. These alterations include oncogenic mutations known to be resistance mechanisms to CDK4/6 inhibitors and endocrine therapy: *AKT1* (PI3K/AKT/mTOR pathway), *NF1* (MAPK pathway), *ERBB2* (RTK pathway), *RB1* (cell cycle), and *ESR1* (ER pathway) (Fig. 2). Our analysis additionally identified novel plausible mechanisms of resistance in the PI3K/AKT/mTOR and MAPK pathways: an activating *MTOR*^T1977R^ mutation, an activating double *PIK3CA*^E545K,G1007R^ mutation, and an activating *BRAF*^V600E^ mutation. An in-depth look at the alterations co-occurring with these potential resistance mechanisms showed some possible confounder alterations but was overall supportive of their proposed role as CDK4/6i-resistance drivers.

Through joint analysis of genomic and transcriptomic tumor data, we discovered a high degree of consistency between the presence of known genomic resistance mechanisms (mutations, high-grade CNAs) and their associated transcriptomic features (gene expression levels, transcriptional signature activities, molecular subtype) (Fig. 3). In particular, we found that all biopsies with activating mutations in *ESR1* showed a concordant high activity in ER pathway signatures, and we also saw an analogous but less strong effect for activating mutations in the PI3K/AKT/mTOR pathway (*AKT1* or *PIK3CA*) and the RTK/MAPK pathway (*ERBB2* or *BRAF*) (Fig. 3A).

The consistency between genomic resistance mechanisms and their associated transcriptomic features suggested some of these features could be used to identify the genes or pathways driving CDK4/6i and endocrine therapy resistance in tumors, particularly in those with no genomic data or no clear genomic drivers. Using the clinical, genomic, and transcriptomic features in their baseline tumors, we identified known or plausible resistance mechanisms to the prior CDK4/6i and anti-ER therapies for 22 out of 23 patients. This included 5 patients in which the only mechanism identified was a transcriptomic feature: 4 patients with no genomic data, and 1 patient with genomic data but no clear genomic drivers. These results highlight how genomic and transcriptomic data complement each other and can identify putative drug resistance drivers in tumors.

### Potential association between genomic alterations driving resistance and transcriptomic-based molecular subtype

A notable novel finding from our genomic/transcriptomic analysis was that activating *ESR1* mutations were exclusive to samples with a Luminal A or B subtype and that activating *ERBB2* or *BRAF* mutations were exclusive to samples with a HER2-E subtype, making these groups mutually exclusive (Fig. 3B). This mutual exclusivity between *ESR1* and RTK/MAPK pathway mutations is consistent with previous observations in HR+/HER2-MBC (Razavi et al. 2018; Nayar et al. 2019; Mao et al. 2020), but to our knowledge, this is the first time the presence of these oncogenic mutations has been linked to the PAM50 molecular subtype of a tumor.

The association between ER and RTK/MAPK alterations and the PAM50 subtype brings up the possibility that some RTK/MAPK oncogenic drivers in HR+/HER2-MBC induce resistance to endocrine therapy by switching the tumor towards an ER pathway-independent transcriptional state. In other words, the resistance mechanism class underlying some RTK/MAPK oncogenic drivers might be closer to “ Pathway Indifference” than “ Pathway Bypass” of the ER pathway (Konieczkowski, Johannessen, and Garraway 2018; Vasan, Baselga, and Hyman 2019). In this view, just as HR+/HER2-BC is endocrine therapy sensitive and most-often in a Luminal A/B transcriptional state and HR-/HER2+ BC is endocrine therapy insensitive and most-often in a HER2-E transcriptional state, certain RTK/MAPK oncogenic drivers might be able push HR+/HER2-MBC tumors to a HER2-E transcriptional state that is endocrine therapy insensitive.

Notably missing from the association between ER and RTK/MAPK mutations were a subset of known RTK/MAPK alterations (*NF1* loss-of-function mutations, *FGFR1* activating mutations and high amplifications). In our case, all samples with these RTK/MAPK alterations also had an activating *ESR1* mutation (Supplemental Fig. S2B) and did not show high activity in RTK pathway signatures (Supplemental Fig. S2A). Co-occurrence of *FGFR1* high amplifications and activating *ESR1* mutations is relatively common (2 cases out of 5 with an *FGFR1* high amplification in our prior study (S. A. Wander et al. 2020)), but co-occurrence *NF1* loss-of-function mutations or *FGFR1* activating mutations and activating *ESR1* mutations are rare (3 cases out of 91 with these *NF1* or *FGFR1* mutations from two recent MSK-IMPACT studies (Q. Li et al. 2022; Razavi et al. 2018)). The observed link between molecular subtype and activating mutations in *ESR1* (Luminal A/B) and *ERBB2/BRAF* (HER2-E) hints at the following possibility for the cases where there are both *ESR1* and RTK/MAPK oncogenic alterations (e.g. in *NF1* and *FGFR1*): *ESR1* mutations drive the tumor towards an ER pathway-dependent transcriptional state that cannot be overcome by RTK/MAPK alterations, resulting in a low RTK pathway signature activity and a Luminal A or B subtype.

### The complexity of the process of acquired resistance to CDK4/6i therapy revealed by tumor evolutionary analysis

Tumor evolutionary analysis of pre- and post-CDK4/6i samples identified acquired genomic alterations in multiple resistance pathways (Figs. 5 and 6). Notable cases include an *ERBB2* amplification in a tumor with a pre-existing *ERBB2*^L869R^ activating mutation, tumors in which the only acquired or enriched alterations were in the ER pathway (*ESR1*^*H524L*^, *ESR1*^D538G^, *GATA3*^*-412fs*^), and a clonal *BRAF*^V600E^ activating mutation in a tumor with concurrent ER loss (possibly the first such mutation in the context of HR+/HER2-MBC; no activating *BRAF* mutations were reported in (S. A. Wander et al. 2020; Z. Li et al. 2018; Bertucci et al. 2019; Nayar et al. 2019; Mao et al. 2020) and the two reported in (Razavi et al. 2018) were subclonal). For two patients, we identified co-existing tumor lineages with distinct driver mutations, one dominating the tumor sample and one the ctDNA sample. In one of these patients, each tumor lineage acquired a genomic alteration in HER2 (*ERBB2* amplification, *ERBB2*^L755S^ mutation), demonstrating convergent evolution of *ERBB2* activation following CDK4/6i therapy and possibly the first reported case of its kind (Fig. 5A).

The distinct co-existing tumor lineages from tumor and blood biopsy samples illustrate the complexity of the process of acquired drug resistance across a patient’s tumors. This complexity is particularly evident in the convergent (*ERBB2* activation) and divergent (*ESR1* or *MTOR* activation) paths to resistance identified. Genomic analysis of co-existing tumor lineages provides a unique window into the evolutionary dynamics of tumor drug resistance, and can help identify high-confidence resistance mechanisms, as we did here for *ERBB2* activation and like prior work has done for various targeted therapies (S. A. Wander et al. 2020; Juric et al. 2015). In our work, this was only possible because of the availability of WES for both tumor and blood biopsy samples, and the existence of these distinct co-existing lineages was not uncommon (18%, 2/11 of cases with both sample types). These results highlight the drug resistance insights that can be gained from collecting and analyzing joint tumor and blood biopsy samples.

### mTOR pathway activation as a molecular correlate of response to combined palbociclib, exemestane, and everolimus

Our genomic and transcriptomic analysis of baseline trial samples uncovered hints that clinical benefit to combined ER, CDK4/6, and MTOR inhibition is correlated with activation of the mTOR pathway (Fig. 7). In particular, we found higher mTORC1 pathway activity and PI3K/AKT/mTOR mutations (including an activating *MTOR* mutation) in clinical benefit-associated samples. We also found that our results were inconclusive in terms of whether *AKT1*^*E17K*^ mutations are a marker for response to triplet therapy, as one would hypothesize based on everolimus targeting the PI3K/AKT/mTOR pathway. This was because of the four patients with *AKT1*^*E17K*^ -mutant tumors, two discontinued therapy due to treatment toxicity, while for the remaining cases one derived clinical benefit and one did not.

Our findings that activation of the mTOR pathway correlates with response to triple therapy are only hypothesis-generating given the limited number of patients that derived clinical benefit. However, they are consistent with recent work finding a more durable response to everolimus-containing therapy in *AKT1*^*E17K*^ vs *AKT1*–wild-type ER+ MBC, with the largest effect seen in those patients with *AKT1*^*E17K*^ and an additional PI3K pathway mutation (Smyth et al. 2020). More definitive answers could come from comprehensive genomic analyses of other triplet therapy trials like TRINITI-1 (Bardia et al. 2021), IPATunity150 (Oliveira et al. 2022), and others (Layman et al. 2022).

To conclude, our multi-omics analysis revealed novel features associated with CDK4/6 inhibitor resistance. These include concurrent *ERBB2* or *BRAF* mutations and a HER2-enriched subtype, and convergent evolution of *ERBB2* activation. Genomic/transcriptomic correlates of response to everolimus-containing triplet therapy hinted at mTOR pathway activation being associated with clinical benefit. These results illustrate how transcriptome sequencing provides complementary and additional information than genome sequencing, and how integrating genomic and transcriptomic data may help better identify patients likely to respond to CDK4/6 inhibitor therapies.

## Methods

Some of the methods reported follow closely what has been described in recent work by some of us (S. A. Wander et al. 2020; Condorelli et al. 2018). For additional details on the methods, refer to the Supplemental Text.

### Patients and samples

Prior to any study procedures, all patients provided written informed consent to participate in the clinical trial and for research biopsies, blood samples, and sequencing of these samples, as approved by the Dana-Farber/Harvard Cancer Center IRB (DF/HCC Protocols 05-246 and 16-177). Metastatic core biopsies were obtained from patients, and samples were immediately snap-frozen in OCT and stored in −80°C. Archival formalin-fixed, paraffin-embedded (FFPE) blocks of primary tumor samples were also obtained. A blood sample was obtained and whole blood was stored at −80°C. DNA and RNA were extracted from tumors. Germline DNA was extracted from peripheral blood mononuclear cells from whole blood. Cell-free DNA was obtained from plasma for circulating tumor DNA analysis, as described previously (Adalsteinsson et al. 2017). For patient 22, targeted panel sequencing data (OncoPanel) for two tumors was obtained (Garcia et al. 2017).

### Phase I/II clinical trial

The protocol for the phase I/II clinical trial (NCT02871791) is available at www.clinicaltrials.gov, and includes information related to the study location, eligibility, and compounds. The primary objective of phase Ib was to determine the safety and tolerability of the combination regimen of palbociclib, everolimus, and exemestane, and to define the maximum tolerated dose (MTD) and recommended phase 2 dose (RP2D). The primary endpoint of phase IIa was to determine the clinical benefit rate (CBR) (complete response + partial response + stable disease ≥ 24 weeks) of the combination regimen. Eligible participants had histologically or cytologically confirmed HR+/HER2-metastatic breast cancer, were required to have radiological or objective evidence of progression on a CDK4/6 inhibitor in the metastatic setting, and relapse or progression on a non-steroidal aromatase inhibitor. Additional information on the clinical trial methods can be found in the Supplemental Text.

### Clinical annotations

Patient charts were reviewed to determine the sequence of treatments received in the neoadjuvant, adjuvant, and metastatic setting, the treatments’ dates and durations, and the dates of the available biopsy samples. The dates and duration of the prior CDK4/6 inhibitor treatment was obtained for all patients for which exome or transcriptome sequencing passed quality control in at least one sample. The dates and duration of the complete sequence of treatments up until discontinuation of the clinical trial regime were obtained for selected patients with 2 or more tumor or cfDNA samples (Figs. 5 and 6).

### Whole exome sequence data processing and analysis

#### Data processing and quality control

Whole exome sequences for each tissue/blood sample were captured using Illumina technology and the sequencing data processing and analysis was performed using the Picard and Terra pipelines at the Broad Institute. The Picard pipeline (http://picard.sourceforge.net) was used to produce a BAM file with aligned reads. This includes alignment to the GRCh37 human reference sequence using the BWA aligner (H. Li and Durbin 2009) and estimation and recalibration of base quality score with the Genome Analysis Toolkit (GATK) (McKenna et al. 2010).

A custom-made cancer genomics analysis pipeline was used to identify somatic alterations using the Terra platform (https://app.terra.bio/). The CGA WES Characterization pipeline developed at the Broad Institute was used to call, filter and annotate somatic mutations and copy number variation. See supplementary methods for details of the variant calling pipeline used in this study. GATK CNV (Van der Auwera and O’Connor 2020) was used for the generation of accurate relative copy-number profiles from the whole exome sequencing data and reference/alternate read counts at heterozygous SNP sites present in both the normal and tumor samples. After accurate proportional coverage profiles are generated for a sample, Allelic CapSeg tool (Landau et al. 2013) was used to generate a segmented allelic copy ratio profile. Allelic copy number profile and mutational call data were modeled jointly by ABSOLUTE (Carter et al. 2012) to produce purity for the samples, a discrete copy number profile, and compute cancer cell fractions (CCF). Tumor samples which had a purity of 8% or more were used for downstream analysis.

#### Annotating oncogenic mutations

OncoKB (Chakravarty et al. 2017) was used to annotate known oncogenic mutations, identify their effect (e.g. loss of function or gain of function), and if they are known cancer hotspots.

#### Tumor evolutionary analysis

Patients with >1 tumor or cfDNA WES samples collected from different timepoints/locations were used to study tumor evolution and tumor heterogeneity. To properly compare SNVs and indels in samples from the same patient, the union of all mutations called in each patient’s samples were considered. The reference and alternate reads in each patient’s samples were used as input for ABSOLUTE (Carter et al. 2012) to compute cancer cell fractions. The clonal structure and the evolutionary history of the clones (phylogenic tree) was inferred with PhylogicNDT (Leshchiner et al. 2019) using only SNV sites and retaining only clones with at least four mutation and cancer cell fraction of more than 1%.

#### Corrected quantification of copy number, gene deletions, and biallelic inactivations

The inference of gene amplifications, gene deletions, and biallelic inactivations were based on the copy number profile obtained from ABSOLUTE (Carter et al. 2012). To infer biallelic inactivations, mutational events that included both loss of heterozygosity (LOH) and a loss-of-function mutation (LOF) (loss-of-function or likely loss-of-function OncoKB-annotated mutation, or a Nonsense_Mutation, Nonstop_Mutation, Frame_Shift_Del, or Frame_Shift_Del mutation) were used. Gene amplifications and deep deletions were based on the purity corrected measure for the segment containing that gene. Genes in a segment-specific copy number of less than 0.5 were considered deep deletions (Deep DEL). To measure segment-specific copy number amplifications, the genome ploidy was subtracted for each sample to obtain the copy number above ploidy (CNAP). CNAPs of at least 3 are considered as amplifications (AMP); CNAPs above 1.5, but below 3 are considered low amplification (GAIN); CNAPs of at least 6 are considered high amplifications (High AMP), and CNAPs of at least 9 and no more than 100 genes are considered focal high amplification (Focal High AMP). In the figures, AMP or GAIN copy number amplifications were not included for any sample and Deep DELs were not included for cfDNA samples (because of limited concordance between tumor and cfDNA Deep DELs, unlike most other alterations).

### Statistical tests and analysis

Statistical significance of the association between signature activities and oncogenic mutations was measured using a one-sided Mann–Whitney test, and is based on whether the AUC ROC score outperforms a random classifier (Mason and Graham 2002). Statistical significance of the association between the Hallmark signature scores in baseline tumors and clinical benefit was measured using a two-sided Welch’s t-test. Statistical significance of the enrichment of upper or lower quartile Hallmark signature scores in the baseline tumors of patients that derived clinical benefit was measured using a two-sided Fisher exact test. A *P* value of < 0.05 was considered to be statistically significant. All statistical analysis was performed using R (version 4.0.3).

### Transcriptome sequencing data processing and analysis

#### Data processing and quality control

RNA-seq reads were mapped to the human genome (hg19) with STAR aligner (Dobin et al. 2013) with default parameters. Transcriptome quality was assessed using RNA-SeQC 2 (Graubert et al. 2021) and expression quantification was conducted using RSEM (B. Li and Dewey 2011). Samples with <8,000 unique genes were removed from subsequent analysis. Gene expression was measured using log2(TPM+1) values and corrected for tissue type (frozen vs FFPE) using ComBat (Leek et al. 2012). The exception was the analysis in Fig. 7B, in which only baseline (T1) samples from frozen tissue were considered, and uncorrected log2(TPM+1) values were used.

#### Transcriptional signature activity

To calculate the activity of a transcriptional signature or gene set, we used single-sample gene set enrichment analysis (ssGSEA). ssGSEA was performed for all tumor samples using fgsea (Korotkevich et al. 2021) to calculate normalized enrichment scores for Hallmark gene sets from the Molecular Signatures Database (Liberzon et al. 2015) using upper quartile normalized expression values.

#### PAM50 molecular subtype assignment

To assign research-based PAM50 subtypes, expression values were rescaled relative to those of a receptor status-balanced Metastatic Breast Cancer Project (MBCProject) cohort, in which samples were re-sampled to achieve the ER-positive to ER-negative receptor status ratio in the UNC training set, from which the PAM50 subtype centroids were derived (A. Prat and Parker 2020; Parker et al. 2009). genefu was used to call research-based PAM50 subtypes (Gendoo et al. 2016) using the rescaled expression values and spearman correlation to the PAM50 subtype centroids. Samples with a PAM50 centroid correlation <0.10 for each centroid were not assigned a PAM50 subtype (Not Classified, NC).

#### Relative gene expression and transcriptional signature activity

To calculate the quantiles for gene expression and transcriptional signature activity, the receptor status-balanced MBCProject cohort was used as a reference cohort. Upper quartile normalized expression was used for relative gene expression and ssGSEA normalized enrichment scores calculated from upper quartile normalized expression was used for relative transcriptional signature activity.

## Data Availability

Tumor and germline whole-exome sequencing data and RNA sequencing data generated and analyzed for this study are in the process of being deposited in dbGaP and will be available upon publication of the manuscript. Additional data generated in this study including clinical trial data, patient metadata, and tumor exome analysis are available in the Supplementary Data.

## Acknowledgements

We thank all the patients who participated in the clinical trial and provided the samples analyzed in this work, without whom this work would not have been possible. This work was supported by an ASPIRE grant from Pfizer (to S. Tolaney), Saverin Breast Cancer Research Fund (to Dana-Farber/Harvard Cancer Center), NCI Specialized Program of Research Excellence (SPORE) Grant 1P50CA168504 (to Dana-Farber/Harvard Cancer Center), Susan G. Komen CCR15333343 (to N. Wagle), The Breast Cancer Alliance (to N. Wagle), The Cancer Couch Foundation (to N. Wagle), and Twisted Pink (to N. Wagle).

## Supplemental Material

**Supplemental Text. Additional information on the clinical trial and supplemental methods**.

**Supplemental Fig. S1. Genomic landscape of resistance to CDK4/6 inhibitors baseline paired tumor and blood biopsies from the clinical trial**. Comutation plot (CoMut) representing the genomic landscape of baseline paired tumor and blood (ctDNA) biopsies from the clinical trial (*n* = 22 samples from *n* = 11 patients). For 9 out of 11 patients, there were few noteworthy differences between each patient’s tumor and ctDNA samples. For 2 out of 11 patients, patients 22 and 32, there were mutually exclusive clonal driver mutations in cancer genes (*ERBB2, ESR1, MTOR*) between their tumor and ctDNA samples, indicative of co-existing distinct tumor lineages. Clinical parameters and genes included, and the arrangement of genes and samples is the same as in Fig. 2. Biopsies are ordered by treatment duration on triplet therapy. Copy-number alterations and nonsynonymous mutations from selected genes are shown. Genes are arranged based on their pathway and include all genes with 2 or more known oncogenic mutations in the cohort. Clinical parameters shown include trial treatment information (trial treatment duration, clinical benefit and best response by RECIST 1.1, reason for discontinuation of treatment), prior CDK4/6i treatment information (CDK4/6i received, antiestrogen agent used in combination, phenotype based on prior CDK4/6i response), receptor status (biopsy-level, at primary diagnosis, and at metastatic diagnosis), timing of biopsy relative to metastatic diagnosis, and biopsy site. Research-based PAM50 subtype (when RNA-seq data is available) and tumor mutational burden of each biopsy are also shown.

**Supplemental Fig. S2. Genomic and transcriptomic features in genes and pathways associated with CDK4/6i and anti-ER tx resistance**. Expanded version of Fig. 3 for the joint genomic and transcriptomic analysis on all baseline trial tumor biopsies with both WES and RNA-seq and a non-Normal PAM50 subtype (*n* = 12 samples and patients). (A) Comutation plot (CoMut) showing the consistency between the presence of oncogenic alterations and the activity of transcriptional signatures of their associated signaling pathway. Distinct genes and signatures are shown depending on the pathway (ER, PI3K/AKT/mTOR, RTK/MAPK, and P53). (B) CoMut dislplaying the association between the presence of oncogenic mutations in *ERBB2* or *BRAF* and a HER2-enriched subtype, and oncogenic mutations in *ESR1* and a Luminal A or B subtype. The arrangement of samples is the same as in the respective CoMuts in Fig. 3, and additionally includes a CoMut with Cell Cycle pathway genes. Biopsies are ordered based on the combined activity of the pathway signatures (panel A) or their correlation to the HER2-enriched centroid (panel B). For each pathway in panel A, all genes from Fig. 2 are shown. For panel B, all genes from Fig. 2 with at least one genomic alteration in these samples are shown. Clinical features shown in each CoMut are HR and HER2 receptor status, ER percentage by IHC, HER2 IHC score, and biopsy site. Transcriptomic-based features shown in each CoMut are research-based PAM50 and Spearman correlation to each PAM50 centroid (excluding the Normal subtype). Quantiles for transcriptional signature activity and gene expression levels are derived from MBCProject.

**Supplemental Fig. S3. Grouped genomic and transcriptomic features in pathways associated with resistance to CDK4/6 inhibitors and antiestrogen treatment in patient’s tumors**. Companion figure for Fig. 4. The chart captures whether key features associated with resistance to CDK4/6 inhibitors or antiestrogen agents are detected in the trial baseline biopsies of patients (*n* = 23 patients with either WES or RNA-seq of a baseline sample). Features are grouped by activation of key genes and pathways. The feature type encodes the data type in which the feature is detected (gene expression, nonsynonymous mutation, copy number alteration). RNA: RNA-seq-based gene expression; CNA: WES-based high-grade copy number alteration; Mutation: WES-based nonsynonymous mutation.

**Supplemental Table S1. Patient disposition at time of data cutoff for the phase II portion of the clinical trial**.

**Supplemental Table S2. Baseline patient, tumor, and treatment characteristics for the phase II portion of the clinical trial**.

**Supplemental Table S3. Best response by RECIST 1.1 for the phase II portion of the clinical trial**. *Patient 15 – Withdrew from trial prior to any scans. Patient 31 – Physician decision was made after the first cycle that toxicities were unacceptable. ^+^n=31 who were off treatment by date of data cutoff for clinical trial.

**Supplemental Table S4. Adverse events possibly, probably, or definitely related to the study drugs (palbociclib, everolimus, exemestane) for the phase II portion of the clinical trial**.

**Supplemental Table S5. Dose reductions/holds due to toxicity for each drug for the phase II portion of the clinical trial**.

**Supplemental Table S6. Patient-level clinical characteristics of cohort**. Excel file, 2 tabs. (Tab 1) Clinical trial patient-level metadata. (Tab 2) Additional patient-level metadata.

**Supplemental Table S7. Sample-level clinical information of cohort**. Excel file, 2 tabs. (Tab 1) Sample-level metadata for samples with exome or transcriptome sequencing data. (Tab 2) Sample-level metadata for samples with targeted panel sequencing data.

**Supplemental Table S8. Genomic information (mutation and copy number calls) of cohort**. Excel file, 4 tabs. (Tab 1) Single nucleotide variants (SNVs) and Indels from whole-exome sequencing (WES). (Tab 2) Copy number variants (CNVs) from WES. (Tab 3) Mutation calls for tumor samples using a targeted panel. (Tab 4) CNVs for tumor samples using a targeted panel. (Tab 5) Selected oncogenes and tumor suppressor genes for WES CNVs.

**Supplemental Table S9. Clinical, genomic, and transcriptomic features associated with resistance to CDK4/6 inhibitors and antiestrogen treatment used in Figure 4 and Supplemental Figure S3**. Excel file, 2 tabs. (Tab 1) Features used in Figure 4. (Tab 2) Features used in Supplemental Figure S3.

**Supplemental Table S10. Treatment timeline for the 6 patients included in the tumor evolutionary analysis of Figures 5 and 6**. Excel file, 1 tab.

**Supplemental Table S11. Results of comparing the Hallmark signature activity in baseline tumors with or without clinical benefit used in Figure 7**. Excel file, 2 tabs. Normalized enrichment scores from single-sample gene set enrichment analysis are used in the comparisons. *n* = 15 tumor samples from baseline (T1) tumors with fresh frozen tissue and a non-Normal PAM50 were used in the comparisons. (Tab 1) Welch’s t-test (two-sided) results comparing Hallmark signature activity in tumors with or without clinical benefit. Uncorrected log2(TPM+1) expression values were used in the comparisons to avoid any potential biases introduced in the tissue-type corrected expression values. (Tab 2) Fisher exact test (two-sided) results comparing enrichment Hallmark signature activity in the upper or lower quartiles in baseline (T1) tumors with or without clinical benefit. Quantiles are based on the tissue-type corrected log2(TPM+1) expression values using the Metastatic Breast Cancer Project as a reference cohort.

## References

Adalsteinsson, Viktor A., Gavin Ha, Samuel S. Freeman, Atish D. Choudhury, Daniel G. Stover, Heather A. Parsons, Gregory Gydush, et al. 2017. “ Scalable Whole-Exome Sequencing of Cell-Free DNA Reveals High Concordance with Metastatic Tumors.” Nature Communications 8 (1): 1324.

Bardia, Aditya, Sara A. Hurvitz, Angela DeMichele, Amy S. Clark, Amelia Zelnak, Denise A. Yardley, Meghan Karuturi, et al. 2021. “ Phase I/II Trial of Exemestane, Ribociclib, and Everolimus in Women with HR+/HER2-Advanced Breast Cancer after Progression on CDK4/6 Inhibitors (TRINITI-1).” Clinical Cancer Research: An Official Journal of the American Association for Cancer Research 27 (15): 4177–85.

Baselga, José, Mario Campone, Martine Piccart, Howard A. Burris, Hope S. Rugo, Tarek Sahmoud, Shinzaburo Noguchi, et al. 2012. “ Everolimus in Postmenopausal Hormone-Receptor–Positive Advanced Breast Cancer.” The New England Journal of Medicine 366 (6): 520–29.

Bertucci, François, Charlotte K. Y. Ng, Anne Patsouris, Nathalie Droin, Salvatore Piscuoglio, Nadine Carbuccia, Jean Charles Soria, et al. 2019. “ Genomic Characterization of Metastatic Breast Cancers.” Nature 569 (7757): 560–64.

Bose, Ron, Shyam M. Kavuri, Adam C. Searleman, Wei Shen, Dong Shen, Daniel C. Koboldt, John Monsey, et al. 2013. “ Activating HER2 Mutations in HER2 Gene Amplification Negative Breast Cancer.” Cancer Discovery 3 (2): 224–37.

Carter, Scott L., Kristian Cibulskis, Elena Helman, Aaron McKenna, Hui Shen, Travis Zack, Peter W. Laird, et al. 2012. “ Absolute Quantification of Somatic DNA Alterations in Human Cancer.” Nature Biotechnology 30 (5): 413–21.

Chakravarty, Debyani, Jianjiong Gao, Sarah M. Phillips, Ritika Kundra, Hongxin Zhang, Jiaojiao Wang, Julia E. Rudolph, et al. 2017. “ OncoKB: A Precision Oncology Knowledge Base.” JCO Precision Oncology 2017 (July). https://doi.org/10.1200/PO.17.00011.

Condorelli, R., L. Spring, J. O’Shaughnessy, L. Lacroix, C. Bailleux, V. Scott, J. Dubois, et al. 2018. “ Polyclonal RB1 Mutations and Acquired Resistance to CDK 4/6 Inhibitors in Patients with Metastatic Breast Cancer.” Annals of Oncology: Official Journal of the European Society for Medical Oncology / ESMO 29 (3): 640–45.

Cornell, Liam, Seth A. Wander, Tanvi Visal, Nikhil Wagle, and Geoffrey I. Shapiro. 2019. “ MicroRNA-Mediated Suppression of the TGF-β Pathway Confers Transmissible and Reversible CDK4/6 Inhibitor Resistance.” Cell Reports 26 (10): 2667–80.e7.

Costa, Carlotta, Ye Wang, Amy Ly, Yasuyuki Hosono, Ellen Murchie, Charlotte S. Walmsley, Tiffany Huynh, et al. 2020. “ PTEN Loss Mediates Clinical Cross-Resistance to CDK4/6 and PI3Kα Inhibitors in Breast Cancer.” Cancer Discovery 10 (1): 72–85.

Dobin, Alexander, Carrie A. Davis, Felix Schlesinger, Jorg Drenkow, Chris Zaleski, Sonali Jha, Philippe Batut, Mark Chaisson, and Thomas R. Gingeras. 2013. “ STAR: Ultrafast Universal RNA-Seq Aligner.” Bioinformatics 29 (1): 15– 21.

Formisano, Luigi, Yao Lu, Alberto Servetto, Ariella B. Hanker, Valerie M. Jansen, Joshua A. Bauer, Dhivya R. Sudhan, et al. 2019. “ Aberrant FGFR Signaling Mediates Resistance to CDK4/6 Inhibitors in ER+ Breast Cancer.” Nature Communications 10 (1): 1373.

Fox, Emily M., Todd W. Miller, Justin M. Balko, Maria G. Kuba, Violeta Sánchez, R. Adam Smith, Shuying Liu, et al. 2011. “ A Kinome-Wide Screen Identifies the insulin/IGF-I Receptor Pathway as a Mechanism of Escape from Hormone Dependence in Breast Cancer.” Cancer Research 71 (21): 6773–84.

Freeman-Cook, Kevin, Robert L. Hoffman, Nichol Miller, Jonathan Almaden, John Chionis, Qin Zhang, Koleen Eisele, et al. 2021. “ Expanding Control of the Tumor Cell Cycle with a CDK2/4/6 Inhibitor.” Cancer Cell 39 (10): 1404–21.e11.

Garcia, Elizabeth P., Alissa Minkovsky, Yonghui Jia, Matthew D. Ducar, Priyanka Shivdasani, Xin Gong, Azra H. Ligon, et al. 2017. “ Validation of OncoPanel: A Targeted Next-Generation Sequencing Assay for the Detection of Somatic Variants in Cancer.” Archives of Pathology & Laboratory Medicine 141 (6): 751–58.

Gendoo, Deena M. A., Natchar Ratanasirigulchai, Markus S. Schröder, Laia Paré, Joel S. Parker, Aleix Prat, and Benjamin Haibe-Kains. 2016. “ Genefu: An R/Bioconductor Package for Computation of Gene Expression-Based Signatures in Breast Cancer.” Bioinformatics 32 (7): 1097–99.

Grabiner, Brian C., Valentina Nardi, Kivanc Birsoy, Richard Possemato, Kuang Shen, Sumi Sinha, Alexander Jordan, Andrew H. Beck, and David M. Sabatini. 2014. “ A Diverse Array of Cancer-Associated MTOR Mutations Are Hyperactivating and Can Predict Rapamycin Sensitivity.” Cancer Discovery 4 (5): 554–63.

Graubert, Aaron, François Aguet, Arvind Ravi, Kristin G. Ardlie, and Gad Getz. 2021. “ RNA-SeQC 2: Efficient RNA-Seq Quality Control and Quantification for Large Cohorts.” Bioinformatics, March. https://doi.org/10.1093/bioinformatics/btab135.

Hafner, Marc, Caitlin E. Mills, Kartik Subramanian, Chen Chen, Mirra Chung, Sarah A. Boswell, Robert A. Everley, et al. 2019. “ Multiomics Profiling Establishes the Polypharmacology of FDA-Approved CDK4/6 Inhibitors and the Potential for Differential Clinical Activity.” Cell Chemical Biology 26 (8): 1067–80.e8.

Hanker, Ariella B., Dhivya R. Sudhan, and Carlos L. Arteaga. 2020. “ Overcoming Endocrine Resistance in Breast Cancer.” Cancer Cell 37 (4): 496–513.

Hurvitz, Sara A., Amy S. Clark, Hope S. Rugo, Aditya Bardia, Amelia Zelnak, Denise A. Yardley, Meghan Karuturi, et al. 2022. “ Abstract PD13-03: Ribociclib, Everolimus, Exemestane Triplet Therapy in HR+/HER2− Advanced Breast Cancer after Progression on a CDK4/6 Inhibitor: Final Efficacy, Safety, and Biomarker Results from TRINITI-1.” Cancer Research 82 (4_Supplement): PD13–03.

Hyman, David M., Sarina A. Piha-Paul, Helen Won, Jordi Rodon, Cristina Saura, Geoffrey I. Shapiro, Dejan Juric, et al. 2018. “ HER Kinase Inhibition in Patients with HER2- and HER3-Mutant Cancers.” Nature 554 (7691): 189–94.

Juric, Dejan, Pau Castel, Malachi Griffith, Obi L. Griffith, Helen H. Won, Haley Ellis, Saya H. Ebbesen, et al. 2015. “ Convergent Loss of PTEN Leads to Clinical Resistance to a PI(3)Kα Inhibitor.” Nature 518 (7538): 240–44.

Kalinsky, Kevin, Melissa Kate Accordino, Codruta Chiuzan, Prabhjot Singh Mundi, Meghna S. Trivedi, Yelena Novik, Amy Tiersten, et al. 2022. “ A Randomized, Phase II Trial of Fulvestrant or Exemestane with or without Ribociclib after Progression on Anti-Estrogen Therapy plus Cyclin-Dependent Kinase 4/6 Inhibition (CDK 4/6i) in Patients (pts) with Unresectable or Hormone Receptor–positive (HR), HER2-Negative Metastatic Breast Cancer (MBC): MAINTAIN Trial.” Journal of Clinical Oncology. https://doi.org/10.1200/jco.2022.40.17_suppl.lba1004.

Konieczkowski, David J., Cory M. Johannessen, and Levi A. Garraway. 2018. “ A Convergence-Based Framework for Cancer Drug Resistance.” Cancer Cell 33 (5): 801–15.

Korotkevich, Gennady, Vladimir Sukhov, Nikolay Budin, Boris Shpak, Maxim N. Artyomov, and Alexey Sergushichev. 2021. “ Fast Gene Set Enrichment Analysis.” bioRxiv. https://doi.org/10.1101/060012.

Kwiatkowski, David J., Toni K. Choueiri, André P. Fay, Brian I. Rini, Aaron R. Thorner, Guillermo de Velasco, Magdalena E. Tyburczy, et al. 2016. “ Mutations in TSC1, TSC2, and MTOR Are Associated with Response to Rapalogs in Patients with Metastatic Renal Cell Carcinoma.” Clinical Cancer Research: An Official Journal of the American Association for Cancer Research 22 (10): 2445–52.

Landau, Dan A., Scott L. Carter, Petar Stojanov, Aaron McKenna, Kristen Stevenson, Michael S. Lawrence, Carrie Sougnez, et al. 2013. “ Evolution and Impact of Subclonal Mutations in Chronic Lymphocytic Leukemia.” Cell 152 (4): 714–26.

Layman, R., R. Wesolowski, H. Han, and J. M. Specht. 2022. “ Abstract PD13-02: Phase Ib Expansion Study of Gedatolisib in Combination with Palbociclib and Endocrine Therapy in Women with ER+ Metastatic Breast Cancer.” Cancer Research. https://aacrjournals.org/cancerres/article-abstract/82/4_Supplement/PD13-02/681453.

Leek, Jeffrey T., W. Evan Johnson, Hilary S. Parker, Andrew E. Jaffe, and John D. Storey. 2012. “ The Sva Package for Removing Batch Effects and Other Unwanted Variation in High-Throughput Experiments.” Bioinformatics 28 (6): 882–83.

Leshchiner, Ignaty, Dimitri Livitz, Justin F. Gainor, Daniel Rosebrock, Oliver Spiro, Aina Martinez, Edmund Mroz, et al. 2019. “ Comprehensive Analysis of Tumour Initiation, Spatial and Temporal Progression under Multiple Lines of Treatment.” bioRxiv. https://doi.org/10.1101/508127.

Liberzon, Arthur, Chet Birger, Helga Thorvaldsdóttir, Mahmoud Ghandi, Jill P. Mesirov, and Pablo Tamayo. 2015. “ The Molecular Signatures Database (MSigDB) Hallmark Gene Set Collection.” Cell Systems 1 (6): 417–25.

Li, Bo, and Colin N. Dewey. 2011. “ RSEM: Accurate Transcript Quantification from RNA-Seq Data with or without a Reference Genome.” BMC Bioinformatics 12 (August): 323.

Li, Heng, and Richard Durbin. 2009. “ Fast and Accurate Short Read Alignment with Burrows-Wheeler Transform.” Bioinformatics 25 (14): 1754–60.

Li, Qing, Baishan Jiang, Jiaye Guo, Hong Shao, Isabella S. Del Priore, Qing Chang, Rei Kudo, et al. 2022. “ INK4 Tumor Suppressor Proteins Mediate Resistance to CDK4/6 Kinase Inhibitors.” Cancer Discovery 12 (2): 356–71.

Li, Zhiqiang, Pedram Razavi, Qing Li, Weiyi Toy, Bo Liu, Christina Ping, Wilson Hsieh, et al. 2018. “ Loss of the FAT1 Tumor Suppressor Promotes Resistance to CDK4/6 Inhibitors via the Hippo Pathway.” Cancer Cell 34 (6): 893– 905.e8.

Mao, Pingping, Ofir Cohen, Kailey J. Kowalski, Justin G. Kusiel, Jorge E. Buendia-Buendia, Michael S. Cuoco, Pedro Exman, et al. 2020. “ Acquired FGFR and FGF Alterations Confer Resistance to Estrogen Receptor (ER) Targeted Therapy in ER+ Metastatic Breast Cancer.” Clinical Cancer Research: An Official Journal of the American Association for Cancer Research 26 (22): 5974–89.

Mason, S. J., and N. E. Graham. 2002. “ Areas beneath the Relative Operating Characteristics (ROC) and Relative Operating Levels (ROL) Curves: Statistical Significance and Interpretation.” Quarterly Journal of the Royal Meteorological Society 128 (584): 2145–66.

McKenna, Aaron, Matthew Hanna, Eric Banks, Andrey Sivachenko, Kristian Cibulskis, Andrew Kernytsky, Kiran Garimella, et al. 2010. “ The Genome Analysis Toolkit: A MapReduce Framework for Analyzing next-Generation DNA Sequencing Data.” Genome Research 20 (9): 1297–1303.

Nayar, Utthara, Ofir Cohen, Christian Kapstad, Michael S. Cuoco, Adrienne G. Waks, Seth A. Wander, Corrie Painter, et al. 2019. “ Acquired HER2 Mutations in ER+ Metastatic Breast Cancer Confer Resistance to Estrogen Receptor– directed Therapies.” Nature Genetics 51 (2): 207–16.

O’Leary, Ben, Rosalind J. Cutts, Yuan Liu, Sarah Hrebien, Xin Huang, Kerry Fenwick, Fabrice André, et al. 2018. “ The Genetic Landscape and Clonal Evolution of Breast Cancer Resistance to Palbociclib plus Fulvestrant in the PALOMA-3 Trial.” Cancer Discovery 8 (11): 1390–1403.

Oliveira, Mafalda, Aditya Bardia, Sung-Bae Kim, Naoki Niikura, Cristina Hernando, Gustavo Werutsky, Yoland Antill, et al. 2022. “ Abstract P5-16-11: Ipatasertib (ipat) in Combination with Palbociclib (palbo) and Fulvestrant (fulv) in Patients (pts) with Hormone Receptor-Positive (HR+) HER2-Negative Advanced Breast Cancer (aBC).” Cancer Research 82 (4_Supplement): P5–16 – 11–P5 – 16–11.

Parker, Joel S., Michael Mullins, Maggie C. U. Cheang, Samuel Leung, David Voduc, Tammi Vickery, Sherri Davies, et al. 2009. “ Supervised Risk Predictor of Breast Cancer Based on Intrinsic Subtypes.” Journal of Clinical Oncology: Official Journal of the American Society of Clinical Oncology 27 (8): 1160–67.

Prat, Aleix, Anwesha Chaudhury, Nadia Solovieff, Laia Paré, Débora Martinez, Nuria Chic, Olga Martínez-Sáez, et al. 2021. “ Correlative Biomarker Analysis of Intrinsic Subtypes and Efficacy Across the MONALEESA Phase III Studies.” Journal of Clinical Oncology: Official Journal of the American Society of Clinical Oncology 39 (13): 1458–67.

Prat, A., and J. S. Parker. 2020. “ Standardized versus Research-Based PAM50 Intrinsic Subtyping of Breast Cancer.” Clinical & Translational Oncology: Official Publication of the Federation of Spanish Oncology Societies and of the National Cancer Institute of Mexico 22 (6): 953–55.

Razavi, Pedram, Matthew T. Chang, Guotai Xu, Chaitanya Bandlamudi, Dara S. Ross, Neil Vasan, Yanyan Cai, et al. 2018. “ The Genomic Landscape of Endocrine-Resistant Advanced Breast Cancers.” Cancer Cell 34 (3): 427–38.e6.

Sánchez-Guixé, Mònica, Cinta Hierro, José Jiménez, Cristina Viaplana, Guillermo Villacampa, Erika Monelli, Fara Brasó-Maristany, et al. 2022. “ High mRNA Expression Levels Correlate with Response to Selective FGFR Inhibitors in Breast Cancer.” Clinical Cancer Research: An Official Journal of the American Association for Cancer Research 28 (1): 137–49.

Smyth, Lillian M., Qin Zhou, Bastien Nguyen, Celeste Yu, Eva M. Lepisto, Monica Arnedos, Michael J. Hasset, et al. 2020. “ Characteristics and Outcome of AKT1E17K-Mutant Breast Cancer Defined through AACR Project GENIE, a Clinicogenomic Registry.” Cancer Discovery 10 (4): 526–35.

Spring, Laura M., Seth A. Wander, Fabrice Andre, Beverly Moy, Nicholas C. Turner, and Aditya Bardia. 2020. “ Cyclin-Dependent Kinase 4 and 6 Inhibitors for Hormone Receptor-Positive Breast Cancer: Past, Present, and Future.” The Lancet 395 (10226): 817–27.

Turner, Nicholas C., Yuan Liu, Zhou Zhu, Sherene Loi, Marco Colleoni, Sibylle Loibl, Angela DeMichele, et al. 2019. “ Cyclin E1 Expression and Palbociclib Efficacy in Previously Treated Hormone Receptor--Positive Metastatic Breast Cancer.” Journal of Clinical Oncology: Official Journal of the American Society of Clinical Oncology 37 (14): 1169.

Turner, Nicholas, Alex Pearson, Rachel Sharpe, Maryou Lambros, Felipe Geyer, Maria A. Lopez-Garcia, Rachael Natrajan, et al. 2010. “ FGFR1 Amplification Drives Endocrine Therapy Resistance and Is a Therapeutic Target in Breast Cancer.” Cancer Research 70 (5): 2085–94.

Van der Auwera, Geraldine A., and Brian D. O’Connor. 2020. Genomics in the Cloud: Using Docker, GATK, and WDL in Terra. O’Reilly Media.

Vasan, Neil, José Baselga, and David M. Hyman. 2019. “ A View on Drug Resistance in Cancer.” Nature 575 (7782): 299– 309.

Wagle, Nikhil, Brian C. Grabiner, Eliezer M. Van Allen, Eran Hodis, Susanna Jacobus, Jeffrey G. Supko, Michelle Stewart, et al. 2014. “ Activating mTOR Mutations in a Patient with an Extraordinary Response on a Phase I Trial of Everolimus and Pazopanib.” Cancer Discovery 4 (5): 546–53.

Wagle, Nikhil, Corrie Painter, Max Krevalin, Coyin Oh, Kristin Anderka, Katie Larkin, Niall Lennon, et al. 2016. “ The Metastatic Breast Cancer Project: A National Direct-to-Patient Initiative to Accelerate Genomics Research.” Journal of Clinical Oncology: Official Journal of the American Society of Clinical Oncology 34 (15_suppl): LBA1519–LBA1519.

Wander, S. A., O. Cohen, X. Gong, and G. N. Johnson. 2020. “ The Genomic Landscape of Intrinsic and Acquired Resistance to Cyclin-Dependent Kinase 4/6 Inhibitors in Patients with Hormone Receptor–Positive Metastatic Breast Cancer.” Cancer Discovery. https://cancerdiscovery.aacrjournals.org/content/10/8/1174.abstract.

Wander, Seth A., Hyo S. Han, Mark L. Zangardi, Andrzej Niemierko, Veronica Mariotti, Leslie S. L. Kim, Jing Xi, et al. 2021. “ Clinical Outcomes With Abemaciclib After Prior CDK4/6 Inhibitor Progression in Breast Cancer: A Multicenter Experience.” Journal of the National Comprehensive Cancer Network: JNCCN, March, 1–8.

Wander, Seth A., Pingping Mao, Maxwell R. Lloyd, Gabriela N. Johnson, Kailey Kowalski, Utthara Nayar, Lillian M. Guenther, et al. 2021. “ Abstract PD7-08: Igf1r Mediates cdk4/6 Inhibitor (cdk4/6i) Resistance in Tumor Samples and in Cellular Models.” Poster Spotlight Session Abstracts. https://doi.org/10.1158/1538-7445.sabcs20-pd7-08.

Xu, Jianing, Can G. Pham, Steven K. Albanese, Yiyu Dong, Toshinao Oyama, Chung-Han Lee, Vanessa Rodrik-Outmezguine, et al. 2016. “ Mechanistically Distinct Cancer-Associated mTOR Activation Clusters Predict Sensitivity to Rapamycin.” The Journal of Clinical Investigation 126 (9): 3526–40.

Yang, C., Z. Li, T. Bhatt, M. Dickler, D. Giri, M. Scaltriti, J. Baselga, N. Rosen, and S. Chandarlapaty. 2017. “ Acquired CDK6 Amplification Promotes Breast Cancer Resistance to CDK4/6 Inhibitors and Loss of ER Signaling and Dependence.” Oncogene 36 (16): 2255–64.

