## Supplemental Text for "Genomic and Transcriptomic Determinants of Resistance to CDK4/6 Inhibitors and Response to Combined Exemestane plus Everolimus and Palbociclib in Patients with Metastatic Hormone Receptor Positive Breast Cancer"

### Supplemental Text. Additional information on the clinical trial and supplemental methods.

#### Additional information on the clinical trial

##### *Methods*

**Study Design and Patient Population.** Eligible participants had histologically or cytologically confirmed HR+/HER2- MBC. Participants enrolled in the phase Ib portion of the study were required to have evaluable disease, whereas participants enrolled in the phase IIa portion were required to have measurable disease per RECIST 1.1 (Eisenhauer et al. 2009). Participants were also required to have radiological or objective evidence of progression on a CDK4/6 inhibitor in the metastatic setting and relapse or progression on a non-steroidal aromatase inhibitor (NSAI) (defined as either relapse within 12 months after completing adjuvant NSAI or progression through a NSAI for metastatic or locally advanced HR+ breast cancer). Any number of prior endocrine therapies were allowed, as long as none were exemestane-based. Up to one prior line of chemotherapy was allowed. Participants were required to be at least 18 years old with an ECOG PS ≤ 2. In addition, participants enrolled in the phase IIa portion of the study were required to undergo a research biopsy at baseline (before treatment initiation) and at the time of disease progression. Participants were also required to provide a single research blood sample before the initiation of therapy. Participants with demonstrated intolerance to 125 mg palbociclib were ineligible. Additional exclusionary criteria included prior treatment with any mTOR inhibitor or concurrent treatment with other investigational agents.

**Procedures.** Palbociclib was administered orally, once daily for 21 consecutive days, followed by a 7-day rest (28-day cycle). Everolimus and exemestane were administered orally, once daily for 28 consecutive days (28-day cycle). During phase Ib, participants were treated with increasing/decreasing doses of palbociclib and everolimus to establish the MTD/RP2D for both drugs in the context of the P-E-E combination. The starting dose of palbociclib was 100 mg, which was increased to 125 mg. The starting dose of everolimus was 5 mg, which was increased to 10 mg. Only one of the two study drugs was escalated/de-escalated at a time. If patients developed toxicity to 5 mg everolimus, de-escalation to 2.5 mg was allowed. Palbociclib doses below 100 mg were not explored. Exemestane was maintained at 25 mg. Patients were observed for dose-limiting toxicity (DLT) events, which were defined as adverse events or abnormal laboratory values with a reasonably possible relationship to the study medication(s).

The RP2D for the combination was determined to be palbociclib 100 mg + everolimus 5 mg + exemestane 25 mg (Barroso-Sousa et al. 2018). This dose was used throughout phase IIa. During phase IIa, patients were evaluated for response every 8 weeks according to RECIST 1.1 criteria (Eisenhauer et al. 2009). Changes in the largest diameter (unidimensional measurement) of the tumor lesions and the shortest diameter in the case of malignant lymph nodes were used.

**Statistical Considerations.** The primary objective of phase Ib was to determine the safety and tolerability of the combination regimen, and to define the MTD/RP2D. The secondary objectives of this phase were to describe the pharmacokinetics (PK) profile of everolimus and exemestane in the triplet combination and evaluate the potential effect of palbociclib on the PK profile of everolimus. For this portion of the study, participants proceeded in dose escalation following the 3 + 3 rule. Treatment-related toxicities were summarized by maximum grade and by term using CTCAE version 4.0 and reported with 90% binomial exact confidence intervals.

The primary endpoint of phase IIa was to determine the CBR (CR + PR + SD ≥ 24 weeks) of the P-E-E combination. Based on the results of the BOLERO-2 trial (Beaver and Park 2012), we estimated that the true CBR would be 40%. Thus, we determined that a CBR of ≥ 65% would indicate that the P-E-E regimen is worthy of further study. Recruitment of at least 29 participants would provide 90% power to test the hypothesis (one-sided alpha of 0.1 and type II error of 0.1). In order to account for a 10% drop out rate, a total of 32 participants were recruited over the course of two and a half years. Up to an additional six months of follow-up was required on the last participant accrued to observe response after the final cycle of protocol therapy, for a total study duration of 3 years.

Secondary endpoints included objective response rate (ORR), disease control rate (DCR), duration of response (DOR), and PFS according to investigator assessment. These metrics were reported with 90% confidence intervals.

**Compliance with Ethical Standards.** The study was conducted in accordance with the International Conference on Harmonization Good Clinical Practice Standards and the Declaration of Helsinki. Institutional review board (IRB) approval was obtained at Dana-Farber/Harvard Cancer Center (DF/HCC). The DF/HCC Data and Safety Monitoring Committee (DSMC), which is composed of clinical specialists with experience in oncology and who had no direct relationship with the study, reviewed and monitored toxicity and accrual data from the study. Information that raised questions or concerns was addressed with the overall PI and study team. Participants provided written informed consent prior to the performance of any protocol specific procedures or assessments.

#### *Results*

**Phase Ib**. *MTD/RP2D.* A total of 9 patients were recruited into the phase Ib portion of the study. At the starting dose of palbociclib (100 mg), 1 out of 3 patients experienced a DLT (grade 3 neutropenia and grade 2 mucositis). Subsequently, 3 additional patients were initiated at 100 mg, and none experienced a DLT. Palbociclib was increased to 125 mg, and all 3 patients had a DLT (grade 3 neutropenia). Thus, 100 mg palbociclib was declared the MTD. The RP2D for phase IIa was 100 mg palbociclib + 5 mg everolimus + 25 mg exemestane (Barroso-Sousa et al. 2018).

**Phase IIa.** *Patient Characteristics.* A total of 32 patients were recruited into phase IIa of the study between May 26, 2017 and June 26, 2019. Patient disposition at the time of data cutoff (Jan 3, 2021) is reported in Supplemental Table S1. All study participants were female, with a median age of 55.5 years (range: 36-73). Most of the participants were initially diagnosed with stage I-III breast cancer and had a disease-free interval longer than two years. However, a substantial percentage of patients (31.2%) had been diagnosed with *de novo* metastatic breast cancer. Bone (87.5%), liver (81.3%) and CNS + lung + liver (84.4%) were the most common sites of metastases (Supplemental Table S2).

The majority of the participants (62.5%) had not received prior chemotherapy for metastatic disease. Among patients who had received prior chemotherapy in the metastatic setting, capecitabine was the most commonly used agent (21.9%). Nearly all the patients (96.9%) had received at least one prior line of hormonal therapy for metastatic or recurrent disease. In addition, all participants had been treated with a prior CDK4/6 inhibitor; 30 patients (93.8%) had received prior palbociclib for metastatic disease, whereas the remaining 2 patients had received abemaciclib (Supplemental Table S2).

*Efficacy.* After a median follow-up of 23.7 months (Supplemental Table S2), 6 patients (18.8%) had SD ≥ 24 weeks, 18 patients (56.3%) had SD ≥ 12 weeks, and 8 patients (25.0%) had PD as the best response. None of the patients had a confirmed complete or partial response to therapy. The CBR was 18.8% (Supplemental Table S3). The median PFS was 3.94 months (95% CI: 3.68-9.63) (Fig. 1C), and the median OS was 24.7 months (95% CI: 20.6 – NA) (Fig. 1D).

*Safety.* The most common all-grade adverse events related to study treatment were neutropenia (90.6%), oral mucositis (53.1%), thrombocytopenia (28.1%), and fatigue (25.0%). A total of 25 patients (78.1%) experienced neutropenia of grade 3 or higher (Supplemental Table S4). Most treatment-related adverse events were likely caused by palbociclib and/or everolimus in this combination regimen (Supplemental Table S4).

25 patients (78.1%) had a dose hold of palbociclib due to toxicity, and 16 patients (50.0%) had at least one dose reduction. Furthermore, 22 patients (68.8%) had a dose hold of everolimus, and 13 (40.6%) had at least one dose reduction due to toxicity. Exemestane was held in 2 patients (6.3%) due to toxicity, and no participants required a dose reduction of exemestane (Supplemental Table S5).

### Supplemental methods

#### *Ultra Low Pass Whole Genome Sequence data processing and quality control for blood biopsies*

Each of the obtained cfDNA samples are subjected to ultra low coverage whole genome sequencing to estimate the tumor content (using ichorCNA) for each sample (Adalsteinsson et al. 2017). Samples with >= 5% estimated tumor fraction were then submitted for whole exome sequencing.

#### *Somatic alterations assessment*

A custom made cancer genomics analysis pipeline was used to identify somatic alterations using the Terra platform (https://app.terra.bio/). We have utilized the CGA WES Characterization pipeline developed at the Broad Institute to call, filter and annotate somatic mutations and copy number variation (available in the Terra platform public workspace *broad-fc-getzlab-workflows/CGA_WES_Characterization_OpenAccess*). The pipeline employs the following tools: MuTect (Cibulskis et al. 2013), ContEst (Cibulskis et al. 2011), Strelka (Saunders et al. 2012), Orientation Bias Filter (Costello et al. 2013), DeTiN (Taylor-Weiner et al. 2018), AllelicCapSeg (Landau et al. 2013), MAFPoNFilter (Lawrence et al. 2014), BLAT realignment filter, ABSOLUTE (Carter et al. 2012), GATK (McKenna et al. 2010), GATK CNV (Van der Auwera and O'Connor 2020), Picard Tools *CrosscheckFingerprints* and *CollectMultipleMetrics* (https://broadinstitute.github.io/picard/), Variant Effect Predictor (Shamsani et al. 2019), and Oncotator (Ramos et al. 2015). To annotate known oncogenic mutations, the OncoKB (Chakravarty et al. 2017) annotator was used (https://github.com/oncokb/oncokb-annotator).

#### *Tumor mutational burden*

Tumor mutational burden was calculated as the total number of mutations (non-synonymous) detected for a given sample divided by the length of the total genomic target region captured with WES.

#### *Biopsy phenotypes to prior CDK4/6i response*

A phenotype of response to the prior CDK4/6i treatment was assigned to each baseline tumor or cfDNA samples following (Wander et al. 2020), and using duration of treatment as a proxy for treatment response. All baseline samples for which sequencing data passed quality control were taken after progression on the prior CDK4/6 inhibitor. Thus, these post-CDK4/6i samples were assigned an acquired resistance phenotype if time on the prior CDK4/6i treatment was >6 months and an intrinsic resistance phenotype otherwise.

#### *Correcting for expression differences in FFPE and frozen tissue*

In our cohort, sequencing on baseline (T1) samples was performed almost exclusively on frozen tissue while sequencing on all archival samples (A1) and one baseline sample (20_T1) was performed on FFPE tissue. The MBCProject, which we use as a reference cohort, consists of only FFPE tissue. In order to compare the expression of the frozen samples to that of the FFPE samples (both the archival and the MBCProject samples), the log2(TPM+1) expression values of the frozen samples were corrected using ComBat in the sva package (Leek et al. 2012). In all the transcriptomic analyses, the corrected log2(TPM+1) expression values were used. The only exception was the Hallmark signature analysis in Fig. 7B and Tab 1 in Supplemental Table S11, where only baseline (T1) samples from frozen tissue were considered ($n$ = 15 tumor samples, which excludes 2 samples with a Normal PAM50, and 1 FFPE tissue sample) and the uncorrected log2(TPM+1) expression values were used to avoid any potential biases introduced in the tissue-type corrected expression values.

#### *DNA and RNA extraction for tumors and whole blood*

DNA extraction was performed as described previously (48). For whole blood, DNA was extracted using magnetic bead–based chemistry in conjunction with the Chemagic MSM I instrument (Perkin Elmer) or the QIAsymphony SP instrument (Qiagen). Following red blood cell lysis, magnetic beads bound to the DNA and were removed from solution using electromagnetized rods. Several wash steps followed to eliminate cell debris and protein residue from DNA bound to the magnetic beads. DNA was then eluted in TE buffer. For frozen tumor tissue, DNA and RNA were extracted simultaneously from a single frozen tissue or cell pellet sample using the AllPrep DNA/RNA Kit (Qiagen). For FFPE tumor tissues, DNA and RNA were extracted simultaneously using Qiagen's AllPrep DNA/RNA FFPE Kit. All DNA was quantified using Picogreen.

##### *DNA extraction for cfDNA from whole blood*

Blood tubes were centrifuged at 1900 g for 10 minutes and plasma was transferred to second tube before further centrifugation at 15000 g for 10 minutes. Supernatant plasma was stored at -80°C until cfDNA extraction. cfDNA was extracted using the QIAsymphony DSP Circulating DNA Kit according to the manufacturer's instructions, with 6.3 mL of plasma as input and with a 60 µL DNA elution (Qiagen, 2017).

##### *Library construction (exomes)*

**Tumors and whole blood.** DNA libraries for massively parallel sequencing were generated as described previously (Fisher et al. 2011) with the following modifications: the initial genomic DNA input into the shearing step was reduced from 3 µg to 10–100 ng in 50 µL of solution. For adapter ligation, Illumina paired-end adapters were replaced with palindromic forked adapters (purchased from Integrated DNA Technologies) with unique dual indexed 8 base index molecular barcode sequences included in the adapter sequence to facilitate downstream pooling. Kapa HyperPrep reagents in 96-reaction kit format were used for end repair/A-tailing, adapter ligation, and library enrichment PCR. In addition, during the post-enrichment solid-phase reversible immobilization (SPRI) bead cleanup, elution volume was reduced to 30 µL to maximize library concentration, and a vortexing step was added to maximize the amount of template eluted.

**cfDNA.** Initial DNA input was normalized to be within the range of 25-52.5 ng in 50 µL of TE buffer (10 mM Tris HCl 1 mM EDTA, pH 8.0) according to picogreen quantification. Library preparation was performed using a commercially available kit provided by KAPA Biosystems (KAPA HyperPrep Kit with Library Amplification product KK8504) and IDT's duplex UMI adapters. Unique 8-base dual index sequences embedded within the p5 and p7 primers (purchased from IDT) were added during PCR. Enzymatic clean-ups were performed using Beckman Coultier AMPure XP beads with elution volumes reduced to 30 µL to maximize library concentration. Library quantification was performed using the Invitrogen Quant-It broad range dsDNA quantification assay kit (Thermo Scientific Catalog: Q33130) with a 1:200 PicoGreen dilution. Following quantification, each library is normalized to a concentration of 25 ng/µL, using Tris-HCl, 10mM, pH 8.0.

##### *Solution-phase hybrid selection (exomes)*

After library construction, hybridization and capture were performed using the relevant components of Illumina's Nextera Rapid Capture Exome Kit or TruSeq Rapid Exome Kit following the manufacturer's suggested protocol, with the following exceptions: first, all libraries within a library construction plate were pooled prior to hybridization. Second, the Midi plate from Illumina's Exome Kit was replaced with a skirted PCR plate to facilitate automation. All hybridization and capture steps were automated on the Agilent Bravo liquid handling system.

##### *Preparation of libraries for cluster amplification and sequencing (exomes)*

After post-capture enrichment, library pools were then quantified using quantitative PCR (KAPA Biosystems) with probes specific to the ends of the adapters; this assay was automated using Agilent's Bravo liquid handling platform. Based on qPCR quantification, libraries were normalized to 2 nM, then denatured using 0.1 or 0.2 N NaOH on the Hamilton Starlet.

##### *Cluster amplification and sequencing (exomes)*

**Tumors and whole blood.** Cluster amplification of denatured templates was performed according to the manufacturer's protocol (Illumina) using HiSeq 2500 Rapid Run v1/v2, HiSeq 2500 High Output v4, HiSeq 4000 v1, or exclusion amplification cluster chemistry, and HiSeq 2500 (Rapid or High Output), HiSeq 4000, or HiSeq X flowcells. Flowcells were sequenced on HiSeq 2500 using v1 (Rapid Run flowcells) or v4 (High Output flowcells) Sequencing-by-Synthesis chemistry, v1 Sequencing-by-Synthesis chemistry for HiSeq 4000 flowcells, or v2.5 Sequencing-by-Synthesis chemistry for HiSeq X flowcells. The flowcells were then analyzed using RTA v.1.18.64 or later. Each pool of whole exome libraries was run on paired 76 bp runs, reading the dual-indexed sequences to identify molecular indices and sequenced across the number of lanes needed to meet coverage for all libraries in the pool.

**cfDNA.** Cluster amplification of library pools was performed according to the manufacturer's protocol (Illumina) using Exclusion Amplification cluster chemistry and HiSeq X flowcells. Flowcells were sequenced on v2 Sequencing-by-Synthesis chemistry for HiSeq X flowcells. The flowcells are then analyzed using RTA v.2.7.3 or later. Each pool of libraries was run on paired 151 bp runs, reading the dual-indexed sequences to identify molecular indices and sequenced across the number of lanes needed to meet coverage for all libraries in the pool.

##### *cDNA library construction (transcriptomes)*

**Transcriptome capture (FFPE tissue).** Total RNA was assessed for quality using the Caliper LabChip GX2. The percentage of fragments with a size greater than 200nt (DV200) was calculated using software. An aliquot of 200ng of RNA was used as the input for first strand cDNA synthesis using Illumina's TruSeq RNA Access Library Prep Kit. Synthesis of the second strand of cDNA was followed by indexed adapter ligation. Subsequent PCR amplification enriched for adapted fragments. The amplified libraries were quantified using an automated PicoGreen assay.

200 ng of each cDNA library, not including controls, were combined into 4-plex pools. Capture probes that target the exome were added, and hybridized for recommended time. Following hybridization, streptavidin magnetic beads were used to capture the library-bound probes from the previous step. Two wash steps effectively remove any nonspecifically bound products. These same hybridization, capture and wash steps are repeated to assure high specificity. A second round of amplification enriches the captured libraries. After enrichment the libraries were quantified with qPCR using the KAPA Library Quantification Kit for Illumina Sequencing Platforms and then pooled equimolarly. The entire process is in 96-well format and all pipetting is done by either Agilent Bravo or Hamilton Starlet.

**Tru-Seq strand specific large insert RNA sequencing (frozen tissue).** Total RNA was quantified using the Quant-iT™ RiboGreen® RNA Assay Kit and normalized to 5 ng/ul. Following plating, 2 µL of ERCC controls (using a 1:1000 dilution) were spiked into each sample. An aliquot of 200 or 325 ng for each sample was transferred into library preparation which uses an automated variant of the Illumina TruSeq™ Stranded mRNA Sample Preparation Kit. This method preserves strand orientation of the RNA transcript. It uses oligo dT beads to select mRNA from the total RNA sample. It is followed by heat fragmentation and cDNA synthesis from the RNA template. The resultant 400 bp cDNA then goes through dual-indexed library preparation: 'A' base addition, adapter ligation using P7 adapters, and PCR enrichment using P5 adapters. After enrichment the libraries were quantified using Quant-iT PicoGreen (1:200 dilution). After normalizing samples to 5 ng/µL, the set was pooled and quantified using the KAPA Library Quantification Kit for Illumina Sequencing Platforms. The entire process is in 96-well format and all pipetting is done by either Agilent Bravo or Hamilton Starlet.

##### *Illumina sequencing (transcriptomes)*

Pooled libraries were normalized to 2 nM and denatured using 0.1 N NaOH prior to sequencing. Flowcell cluster amplification and sequencing were performed according to the manufacturer's protocols using HiSeq 2000, HiSeq 2500, or NovaSeq 6000. Each run was a 76 (Transcriptome capture) or 101 (Tru-Seq) bp paired-end with an eight-base index barcode read. Data was analyzed using the Broad Picard Pipeline which includes de-multiplexing and data aggregation.

##### *Figure and plot generation and editing*

Plots were generated using R and the packages ggplot2, ggpubr, and ComplexHeatmap. Figures were edited using Microsoft PowerPoint, and Inkscape. The DNA icon in Fig. 1 is an edited version of the C-DNA icon obtained from the open source library of icons bioicons (https://github.com/duerrsimon/bioicons). The C-DNA icon by DBCLS https://togotv.dbcls.jp/en/pics.html is licensed under CC-BY 4.0 Unported https://creativecommons.org/licenses/by/4.0/. The female silhouette in Fig. 1 is an edited version of an icon obtained from the public domain repository of vector clip art Openclipart (https://openclipart.org/).

#### Supplemental Text references


Adalsteinsson, Viktor A., Gavin Ha, Samuel S. Freeman, Atish D. Choudhury, Daniel G. Stover, Heather A. Parsons, Gregory Gydush, et al. 2017. "Scalable Whole-Exome Sequencing of Cell-Free DNA Reveals High Concordance with Metastatic Tumors." *Nature Communications* 8 (1): 1324.

Barroso-Sousa, Romualdo, Hao Guo, William Thomas Barry, Arlindo R. Ferreira, Rebecca Rees, Eric P. Winer, Nikhil Wagle, and Sara M. Tolaney. 2018. "A Phase I Study of Palbociclib (PALBO) plus Everolimus (EVE) and Exemestane (EXE) in Hormone-Receptor Positive (HR )/HER2- Metastatic Breast Cancer (MBC) after Progression on a CDK4/6 Inhibitor (CDK4/6i): Safety, Tolerability and Pharmacokinetic (PK) Analysis." *Journal of Clinical Oncology*. https://doi.org/10.1200/jco.2018.36.15_suppl.1068.

Beaver, Julia A., and Ben H. Park. 2012. "The BOLERO-2 Trial: The Addition of Everolimus to Exemestane in the Treatment of Postmenopausal Hormone Receptor-Positive Advanced Breast Cancer." *Future Oncology* 8 (6): 651–57.

Carter, Scott L., Kristian Cibulskis, Elena Helman, Aaron McKenna, Hui Shen, Travis Zack, Peter W. Laird, et al. 2012. "Absolute Quantification of Somatic DNA Alterations in Human Cancer." *Nature Biotechnology* 30 (5): 413–21.

Chakravarty, Debyani, Jianjiong Gao, Sarah M. Phillips, Ritika Kundra, Hongxin Zhang, Jiaojiao Wang, Julia E. Rudolph, et al. 2017. "OncoKB: A Precision Oncology Knowledge Base." *JCO Precision Oncology* 2017 (July). https://doi.org/10.1200/PO.17.00011.

Cibulskis, Kristian, Michael S. Lawrence, Scott L. Carter, Andrey Sivachenko, David Jaffe, Carrie Sougnez, Stacey Gabriel, Matthew Meyerson, Eric S. Lander, and Gad Getz. 2013. "Sensitive Detection of Somatic Point Mutations in Impure and Heterogeneous Cancer Samples." *Nature Biotechnology* 31 (3): 213–19.

Cibulskis, Kristian, Aaron McKenna, Tim Fennell, Eric Banks, Mark DePristo, and Gad Getz. 2011. "ContEst: Estimating Cross-Contamination of Human Samples in next-Generation Sequencing Data." *Bioinformatics* 27 (18): 2601–2.

Costello, Maura, Trevor J. Pugh, Timothy J. Fennell, Chip Stewart, Lee Lichtenstein, James C. Meldrim, Jennifer L. Fostel, et al. 2013. "Discovery and Characterization of Artifactual Mutations in Deep Coverage Targeted Capture Sequencing Data due to Oxidative DNA Damage during Sample Preparation." *Nucleic Acids Research* 41 (6): e67.

Eisenhauer, E. A., P. Therasse, J. Bogaerts, L. H. Schwartz, D. Sargent, R. Ford, J. Dancey, et al. 2009. "New Response Evaluation Criteria in Solid Tumours: Revised RECIST Guideline (version 1.1)." *European Journal of Cancer* 45 (2): 228–47.

Fisher, Sheila, Andrew Barry, Justin Abreu, Brian Minie, Jillian Nolan, Toni M. Delorey, Geneva Young, et al. 2011. "A Scalable, Fully Automated Process for Construction of Sequence-Ready Human Exome Targeted Capture Libraries." *Genome Biology* 12 (1): R1.

Landau, Dan A., Scott L. Carter, Petar Stojanov, Aaron McKenna, Kristen Stevenson, Michael S. Lawrence, Carrie Sougnez, et al. 2013. "Evolution and Impact of Subclonal Mutations in Chronic Lymphocytic Leukemia." *Cell* 152 (4): 714–26.

Lawrence, Michael S., Petar Stojanov, Craig H. Mermel, James T. Robinson, Levi A. Garraway, Todd R. Golub, Matthew Meyerson, Stacey B. Gabriel, Eric S. Lander, and Gad Getz. 2014. "Discovery and

Saturation Analysis of Cancer Genes across 21 Tumour Types.” *Nature* 505 (7484): 495–501.
Leek, Jeffrey T., W. Evan Johnson, Hilary S. Parker, Andrew E. Jaffe, and John D. Storey. 2012. “The Sva Package for Removing Batch Effects and Other Unwanted Variation in High-Throughput Experiments.” *Bioinformatics* 28 (6): 882–83.
McKenna, Aaron, Matthew Hanna, Eric Banks, Andrey Sivachenko, Kristian Cibulskis, Andrew Kernytsky, Kiran Garimella, et al. 2010. “The Genome Analysis Toolkit: A MapReduce Framework for Analyzing next-Generation DNA Sequencing Data.” *Genome Research* 20 (9): 1297–1303.
Ramos, Alex H., Lee Lichtenstein, Manaswi Gupta, Michael S. Lawrence, Trevor J. Pugh, Gordon Saksena, Matthew Meyerson, and Gad Getz. 2015. “Oncotator: Cancer Variant Annotation Tool.” *Human Mutation* 36 (4): E2423–29.
Saunders, Christopher T., Wendy S. W. Wong, Sajani Swamy, Jennifer Becq, Lisa J. Murray, and R. Keira Cheetham. 2012. “Strelka: Accurate Somatic Small-Variant Calling from Sequenced Tumor–normal Sample Pairs.” *Bioinformatics* 28 (14): 1811–17.
Shamsani, Jannah, Stephen H. Kazakoff, Irina M. Armean, Will McLaren, Michael T. Parsons, Bryony A. Thompson, Tracy A. O’Mara, Sarah E. Hunt, Nicola Waddell, and Amanda B. Spurdle. 2019. “A Plugin for the Ensembl Variant Effect Predictor That Uses MaxEntScan to Predict Variant Spliceogenicity.” *Bioinformatics* 35 (13): 2315–17.
Taylor-Weiner, Amaro, Chip Stewart, Thomas Giordano, Mendy Miller, Mara Rosenberg, Alyssa Macbeth, Niall Lennon, et al. 2018. “DeTiN: Overcoming Tumor-in-Normal Contamination.” *Nature Methods* 15 (7): 531–34.
Van der Auwera, Geraldine A., and Brian D. O’Connor. 2020. *Genomics in the Cloud: Using Docker, GATK, and WDL in Terra*. O’Reilly Media.
Wander, S. A., O. Cohen, X. Gong, and G. N. Johnson. 2020. “The Genomic Landscape of Intrinsic and Acquired Resistance to Cyclin-Dependent Kinase 4/6 Inhibitors in Patients with Hormone Receptor–Positive Metastatic Breast Cancer.” *Cancer Discovery*. https://cancerdiscovery.aacrjournals.org/content/10/8/1174.abstract.
