## Supplemental Figures for "Genomic and Transcriptomic Determinants of Resistance to CDK4/6 Inhibitors and Response to Combined Exemestane plus Everolimus and Palbociclib in Patients with Metastatic Hormone Receptor Positive Breast Cancer"

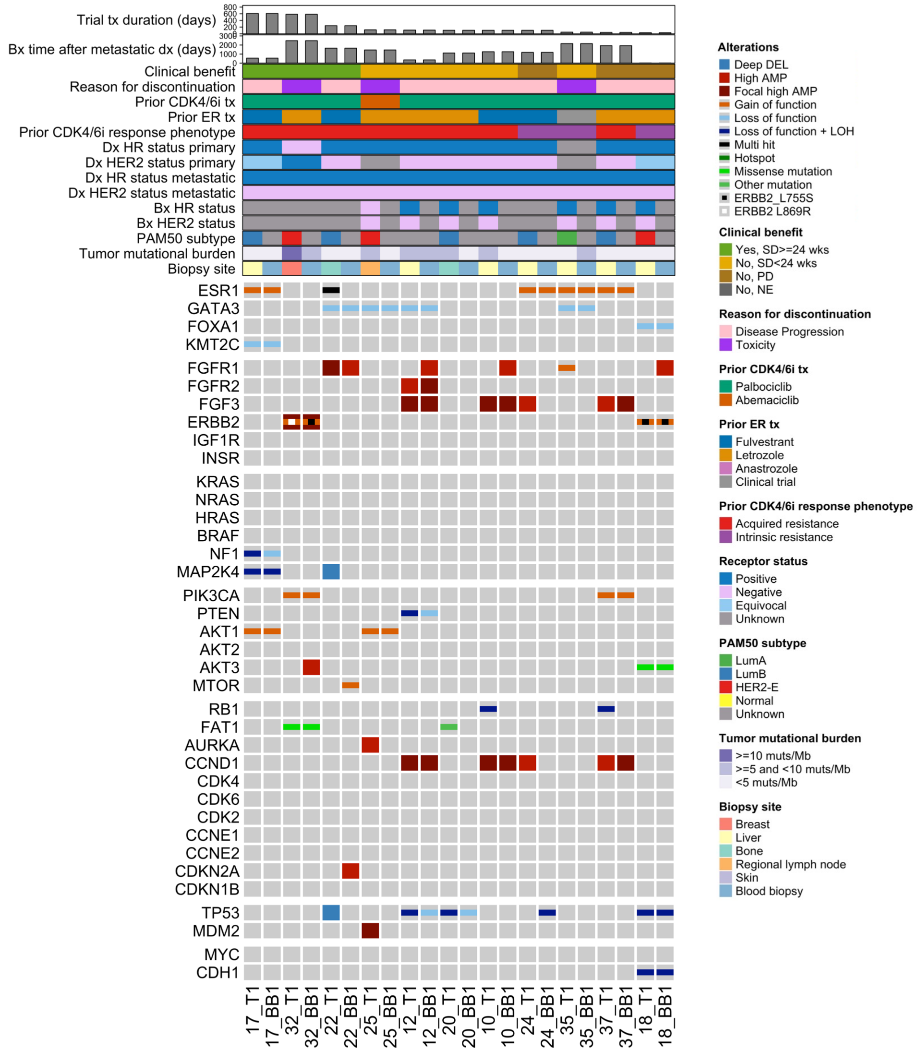


**Supplemental Fig. S1. Genomic landscape of resistance to CDK4/6 inhibitors baseline paired tumor and blood biopsies from the clinical trial.** Comutation plot (CoMut) representing the genomic landscape of baseline paired tumor and blood (ctDNA) biopsies from the clinical trial ($n$ = 22 samples from $n$ = 11 patients). For 9 out of 11 patients, there were few noteworthy differences between each patient's tumor and ctDNA samples. For 2 out of 11 patients, patients 22 and 32, there were mutually exclusive clonal driver mutations in cancer genes (*ERBB2*, *ESR1*, *MTOR*) between their tumor and ctDNA samples, indicative of co-existing distinct tumor lineages. Clinical parameters and genes included, and the arrangement of genes and samples is the same as in Fig. 2. Biopsies are ordered by treatment duration on triplet therapy. Copy-number alterations and nonsynonymous mutations from selected genes are shown. Genes are arranged based on their pathway and include all genes with 2 or more known oncogenic mutations in the cohort. Clinical parameters shown include trial treatment information (trial treatment duration, clinical benefit and best response by RECIST 1.1, reason for discontinuation of treatment), prior CDK4/6i treatment information (CDK4/6i received, antiestrogen agent used in combination, phenotype based on prior CDK4/6i response), receptor status (biopsy-level, at primary diagnosis, and at metastatic diagnosis), timing of biopsy relative to metastatic diagnosis, and biopsy site. Research-based PAM50 subtype (when RNA-seq data is available) and tumor mutational burden of each biopsy are also shown.

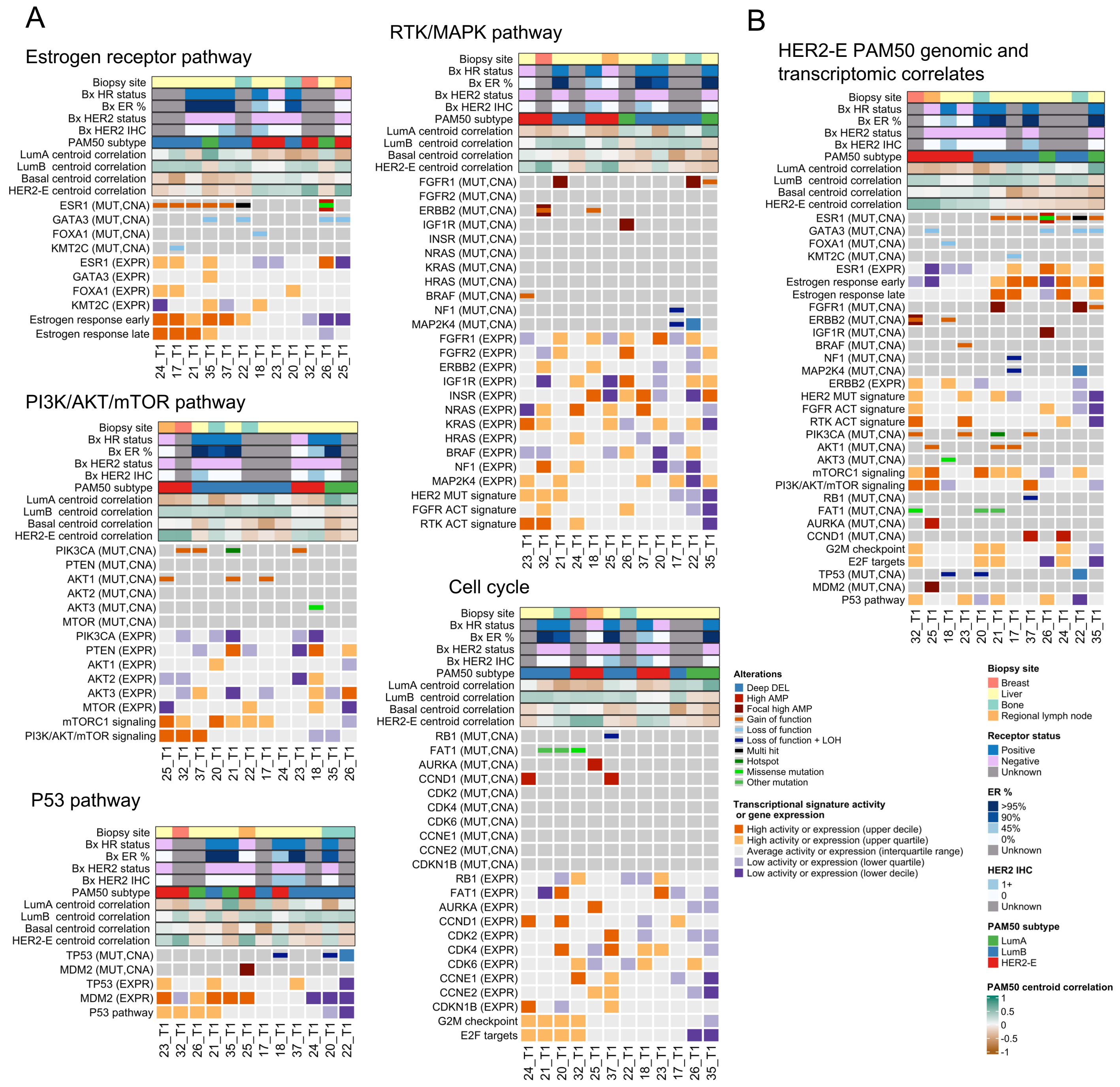


**Supplemental Fig. S2. Genomic and transcriptomic features in genes and pathways associated with CDK4/6i and anti-ER tx resistance.** Expanded version of Fig. 3 for the joint genomic and transcriptomic analysis on all baseline trial tumor biopsies with both WES and RNA-seq and a non-Normal PAM50 subtype (*n* = 12 samples and patients). (A) Comutation plot (CoMut) showing the consistency between the presence of oncogenic alterations and the activity of transcriptional signatures of their associated signaling pathway. Distinct genes and signatures are shown depending on the pathway (ER, PI3K/AKT/mTOR, RTK/MAPK, and P53). (B) CoMut dislplaying the association between the presence of oncogenic mutations in *ERBB2* or *BRAF* and a HER2-enriched subtype, and oncogenic mutations in *ESR1* and a Luminal A or B subtype. The arrangement of samples is the same as in the respective CoMuts in Fig. 2, and additionally includes a CoMut with Cell Cycle pathway genes. Biopsies are ordered based on the combined activity of the pathway signatures (panel A) or their correlation to the HER2-enriched centroid (panel B). For each pathway in panel A, all genes from Fig. 2 are shown. For panel B, all genes from Fig. 2 with at least one genomic alteration in these samples are shown. Clinical features shown in each CoMut are HR and HER2 receptor status, ER percentage by IHC, HER2 IHC score, and biopsy site. Transcriptomic-based features shown in each CoMut are research-based PAM50 and Spearman correlation to each PAM50 centroid (excluding the Normal subtype). Quantiles for transcriptional signature activity and gene expression levels are derived from MBCProject.

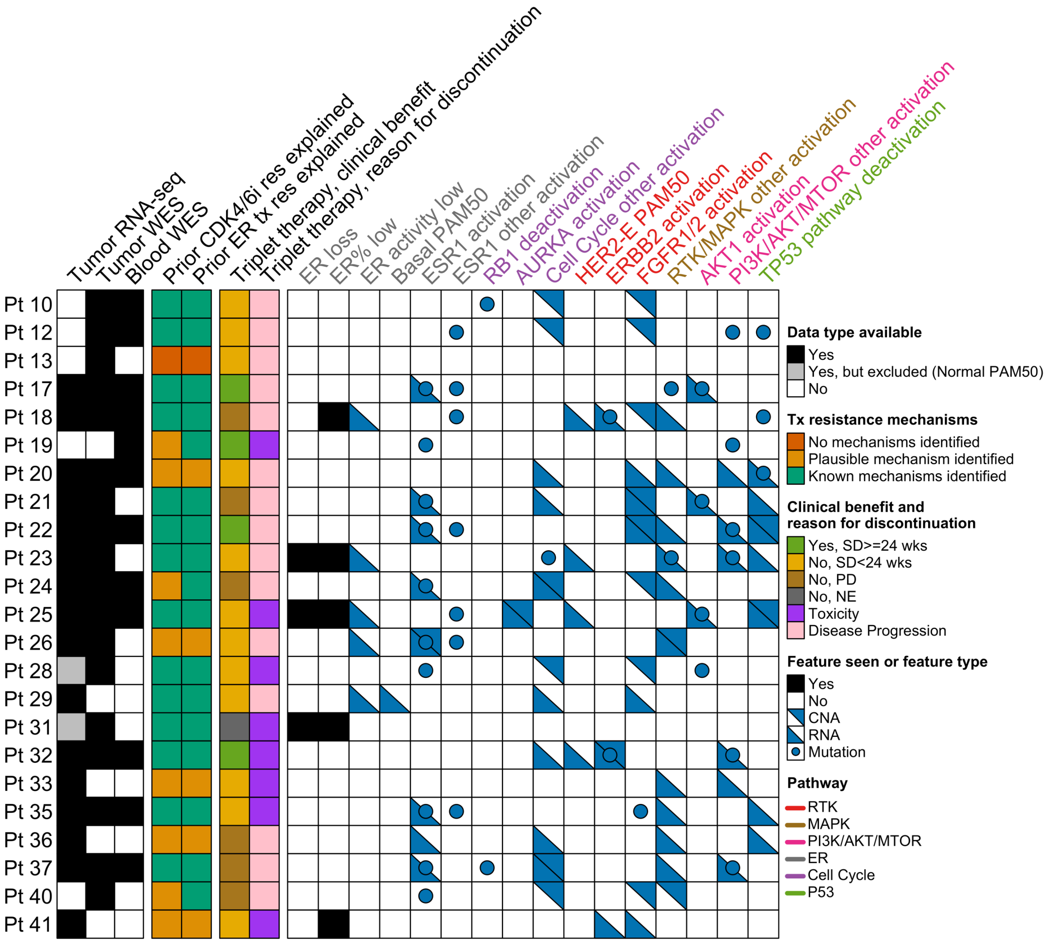


**Supplemental Fig. S3. Grouped genomic and transcriptomic features in pathways associated with resistance to CDK4/6 inhibitors and antiestrogen treatment in patient's tumors.** Companion figure for Fig. 4. The chart captures whether key features associated with resistance to CDK4/6 inhibitors or antiestrogen agents are detected in the trial baseline biopsies of patients (*n* = 23 patients with either WES or RNA-seq of a baseline sample). Features are grouped by activation of key genes and pathways. The feature type encodes the data type in which the feature is detected (gene expression, nonsynonymous mutation, copy number alteration). RNA: RNA-seq-based gene expression; CNA: WES-based high-grade copy number alteration; Mutation: WES-based nonsynonymous mutation.
