## Supplemental Tables S1-S5 for "Genomic and Transcriptomic Determinants of Resistance to CDK4/6 Inhibitors and Response to Combined Exemestane plus Everolimus and Palbociclib in Patients with Metastatic Hormone Receptor Positive Breast Cancer"

**Supplemental Table S1. Patient disposition at time of data cutoff for the phase II portion of the clinical trial.**

| Characteristic | Number of Patients (%) |
|---|---|
| **Number of Patients** | N = 32 |
| **Dates of Registration (Range)** | 05/26/2017-06/26/2019 |
| **Date of Data Cutoff** | 01/03/2021 |
| **Patient disposition at time of data cutoff** | |
| On protocol therapy | 1 (3.1%) |
| Off protocol therapy | 31 (96.9%) |
| **Patient status at time of data cutoff** | |
| Alive | 15 (46.9%) |
| Dead | 16 (50.0%) |
| Lost to follow-up | 1 (3.1%) |
| **Reason for treatment discontinuation** | |
| Progression/relapse based on RECIST 1.1 | 20 (62.5%) |
| Toxicity | 9 (28.1%) |
| Patient withdrew for other reason | 2 (6.3%) |
| **Reason for being off study** | |
| Lost to follow-up | 2 (6.3%) |
| Progression/relapse | 26 (81.2%) |
| Death | 1 (3.1%) |

**Supplemental Table S2. Baseline patient, tumor, and treatment characteristics for the phase II portion of the clinical trial.**

| Characteristic | Number of Patients (%) |
|---|---|
| **Follow-up time in months, median (interquartile range)** | 23.7 (20.1 – 31.8) |
| **Median Age (Range)** | 55.5 (36-73) |
| **Sex** | |
| Female | 32 (100%) |
| **Race** | |
| White | 29 (90.6%) |
| Black | 2 (6.3%) |
| Asian | 1 (3.1%) |
| **Ethnicity** | |
| Hispanic or Latino | 0 (0%) |
| Non-Hispanic | 31 (96.9%) |
| Unknown | 1 (3.1%) |
| **ECOG PS at baseline** | |
| 0 | 27 (84.4%) |
| 1 | 5 (15.6%) |
| **Stage at diagnosis** | |
| I | 7 (21.9%) |
| II | 8 (25%) |
| III | 6 (18.8%) |
| IV | 10 (31.2%) |
| Unknown | 1 (3.1%) |
| **Disease-free interval** | |
| *De novo* metastatic disease | 10 (31.2%) |
| ≤ 2 years | 2 (6.2%) |
| > 2 years | 20 (62.5%) |
| **Sites of disease** | |
| CNS | 0 |
| Lung or pleural effusion | 10 (31.3%) |

| | |
|---|---|
| Liver<br>CNS + lung + liver<br>Bone<br>Breast or chest wall<br>Lymph nodes<br>Soft tissue<br>Other | 26 (81.3%)<br>27 (84.4%)<br>28 (87.5%)<br>4 (12.5%)<br>8 (25.0%)<br>3 (9.4%)<br>6 (18.8%) |
| **Bone only metastases** | 2 (6.3%) |
| **ER, PR, and HER2 status of primary and metastatic tumors** | |
| **Primary tumor**<br>ER+ and/or PR+, HER2-<br>ER- and PR-, HER2+<br>Not done/unknown | <br>23 (71.9%)<br>1 (3.1%)<br>8 (25.0%) |
| **Metastatic/recurrent tumor**<br>ER+ and/or PR+, HER2-<br>ER Status<br>Positive<br>PR Status<br>Low Positive<br>Positive (> 10% Cell Staining)<br>Negative (≤ 10% Cell Staining) | <br>32 (100%)<br><br>32 (100%)<br><br>3 (9.4%)<br>18 (56.3%)<br>11 (34.4%) |
| **Adjuvant or neoadjuvant chemotherapy**<br>Anthracycline<br>Taxane<br>Anthracycline and taxane | <br>7 (21.9%)<br>1 (3.1%)<br>10 (31.3%) |
| **Lines of chemotherapy for metastasis or recurrence**<br>Median (range)<br>0 lines<br>1 line | <br>0 (0-1)<br>20 (62.5%)<br>12 (37.5%) |
| **Lines or hormonal therapy for metastasis or recurrence**<br>Median (range)<br>0 lines<br>1 line<br>2 lines<br>3 or more lines | <br>2 (0-3)<br>1 (3.1%)<br>14 (43.8%)<br>12 (37.5%)<br>5 (15.6%) |
| **Prior metastatic chemotherapy**<br>Anthracycline<br>Taxane<br>Platinum<br>Capecitabine<br>Eribulin<br>Other | <br>1 (3.1%)<br>1 (3.1%)<br>0 (0%)<br>7 (21.9%)<br>2 (6.3%)<br>2 (6.3%) |
| **CDK4/6 inhibitor**<br>Abemaciclib<br>Palbociclib<br>Ribociclib | <br>2 (6.3%)<br>30 (93.8%)<br>0 (0%) |

**Supplemental Table S3. Best response by RECIST 1.1 for the phase II portion of the clinical trial.** *Patient 15 – Withdrew from trial prior to any scans. Patient 31 – Physician decision was made after the first cycle that toxicities were unacceptable. +n=31 who were off treatment by date of data cutoff for clinical trial.

| Response | Number of Patients (%) |
|---|---|
| Confirmed CR | 0 (0%) |
| Confirmed PR | 0 (0%) |
| SD | 22 (68.8%) |
| SD ≥ 12 weeks | 18 (56.3%) |
| SD ≥ 24 weeks | 6 (18.8%) |
| CBR (CR + PR + SD ≥ 24 weeks) | 6 (18.8%) |
| Progressive disease | 8 (25.0%) |
| Not evaluable* | 2 (6.3%) |
| **Median time on treatment in days+** | 111 (28-637) |

**Supplemental Table S4. Adverse events possibly, probably, or definitely related to the study drugs (palbociclib, everolimus, exemestane) for the phase II portion of the clinical trial.**

| Adverse Event (AE) and Drug | Number of Patients (%), all AE | Number of Patients (%), AE Grade 2 | Number of Patients (%), AE Grade ≥3 |
|---|---|---|---|
| **Any of the drugs with ≥ 10% incidence rate** | | | |
| Neutropenia | 29 (90.6%) | 4 (12.5%) | 25 (78.1%) |
| Mucositis – oral | 17 (53.1%) | 12 (37.5%) | 5 (15.6%) |
| Thrombocytopenia | 9 (28.1%) | 6 (18.8%) | 3 (9.4%) |
| Fatigue | 8 (25.0%) | 8 (25.0%) | 0 (0%) |
| Pneumonitis | 7 (21.9%) | 7 (21.9%) | 0 (0%) |
| Anorexia | 6 (18.8%) | 6 (18.8%) | 0 (0%) |
| Aspartate aminotransferase increased | 4 (12.5%) | 1 (3.1%) | 3 (9.4%) |
| **Palbociclib** | | | |
| Neutropenia | 29 (90.6%) | 4 (12.5%) | 25 (78.1%) |
| Mucositis – oral | 16 (50.0%) | 11 (34.4%) | 5 (15.6%) |
| Thrombocytopenia | 8 (25.0%) | 5 (15.6%) | 3 (9.4%) |
| Fatigue | 7 (21.9%) | 7 (21.9%) | 0 (0%) |
| Anorexia | 3 (9.4%) | 3 (9.4%) | 0 (0%) |
| Dehydration | 3 (9.4%) | 2 (6.3%) | 1 (3.1%) |
| Diarrhea | 3 (9.4%) | 2 (6.3%) | 1 (3.1%) |
| Dyspepsia | 2 (6.3%) | 2 (6.3%) | 0 (0%) |
| Gastroesophageal reflux disease | 2 (6.3%) | 2 (6.3%) | 0 (0%) |
| Nausea | 2 (6.3%) | 2 (6.3%) | 0 (0%) |
| Vomiting | 2 (6.3%) | 2 (6.3%) | 0 (0%) |
| Alanine aminotransferase increased | 1 (3.1%) | 0 (0%) | 1 (3.1%) |
| Alkaline phosphatase increased | 1 (3.1%) | 0 (0%) | 1 (3.1%) |
| Anemia | 1 (3.1%) | 1 (3.1%) | 0 (0%) |
| Aspartate aminotransferase increased | 1 (3.1%) | 0 (0%) | 1 (3.1%) |
| Dysgeusia | 1 (3.1%) | 1 (3.1%) | 0 (0%) |

| | | | |
|---|---|---|---|
| Headache | 1 (3.1%) | 1 (3.1%) | 0 (0%) |
| Insomnia | 1 (3.1%) | 1 (3.1%) | 0 (0%) |
| Lymphopenia | 1 (3.1%) | 1 (3.1%) | 0 (0%) |
| Rash – maculo-papular | 1 (3.1%) | 1 (3.1%) | 0 (0%) |
| Sinusitis | 1 (3.1%) | 1 (3.1%) | 0 (0%) |
| Sore throat | 1 (3.1%) | 1 (3.1%) | 0 (0%) |
| Upper respiratory infection | 1 (3.1%) | 1 (3.1%) | 0 (0%) |
| Urinary tract infection | 1 (3.1%) | 1 (3.1%) | 0 (0%) |
| Leukopenia | 1 (3.1%) | 0 (0%) | 1 (3.1%) |
| **Everolimus** | | | |
| Neutropenia | 24 (75.0%) | 4 (12.5%) | 20 (62.5%) |
| Mucositis – oral | 17 (53.1%) | 12 (37.5%) | 5 (15.6%) |
| Pneumonitis | 7 (21.9%) | 7 (21.9%) | 0 (0%) |
| Anorexia | 6 (18.8%) | 6 (18.8%) | 0 (0%) |
| Fatigue | 6 (18.8%) | 6 (18.8%) | 0 (0%) |
| Thrombocytopenia | 5 (15.6%) | 5 (15.6%) | 0 (0%) |
| Aspartate aminotransferase increased | 4 (12.5%) | 1 (3.1%) | 3 (9.4%) |
| Dehydration | 3 (9.4%) | 2 (6.3%) | 1 (3.1%) |
| Diarrhea | 3 (9.4%) | 2 (6.3%) | 1 (3.1%) |
| Alanine aminotransferase increased | 2 (6.3%) | 1 (3.1%) | 1 (3.1%) |
| Alkaline phosphatase increased | 2 (6.3%) | 1 (3.1%) | 1 (3.1%) |
| Anemia | 2 (6.3%) | 1 (3.1%) | 1 (3.1%) |
| Dyspepsia | 2 (6.3%) | 2 (6.3%) | 0 (0%) |
| Nausea | 2 (6.3%) | 2 (6.3%) | 0 (0%) |
| Upper respiratory infection | 2 (6.3%) | 2 (6.3%) | 0 (0%) |
| Vomiting | 2 (6.3%) | 2 (6.3%) | 0 (0%) |
| Abdominal pain | 1 (3.1%) | 1 (3.1%) | 0 (0%) |
| Dysgeusia | 1 (3.1%) | 1 (3.1%) | 0 (0%) |
| Dyspnea | 1 (3.1%) | 1 (3.1%) | 0 (0%) |
| Enterocolitis | 1 (3.1%) | 1 (3.1%) | 0 (0%) |
| Gastric ulcer | 1 (3.1%) | 0 (0%) | 1 (3.1%) |
| Gastrointestinal disorders – Other, specify | 1 (3.1%) | 1 (3.1%) | 0 (0%) |
| Headache | 1 (3.1%) | 1 (3.1%) | 0 (0%) |
| Hyperglycemia | 1 (3.1%) | 1 (3.1%) | 0 (0%) |
| Insomnia | 1 (3.1%) | 1 (3.1%) | 0 (0%) |
| Lung infection | 1 (3.1%) | 1 (3.1%) | 0 (0%) |
| Lymphopenia | 1 (3.1%) | 1 (3.1%) | 0 (0%) |
| Malaise | 1 (3.1%) | 1 (3.1%) | 0 (0%) |
| Pruritis | 1 (3.1%) | 1 (3.1%) | 0 (0%) |
| Rash – acneiform | 1 (3.1%) | 1 (3.1%) | 0 (0%) |
| Rash – maculo-papular | 1 (3.1%) | 1 (3.1%) | 0 (0%) |

| | | | |
|---|---|---|---|
| Rash – pustular | 1 (3.1%) | 1 (3.1%) | 0 (0%) |
| Respiratory, thoracic, and mediastinal disorders | 1 (3.1%) | 1 (3.1%) | 0 (0%) |
| Sore throat | 1 (3.1%) | 1 (3.1%) | 0 (0%) |
| Urinary tract infection | 1 (3.1%) | 1 (3.1%) | 0 (0%) |
| **Exemestane** | | | |
| Anorexia | 1 (3.1%) | 1 (3.1%) | 0 (0%) |
| Aspartate aminotransferase increased | 1 (3.1%) | 1 (3.1%) | 0 (0%) |
| Headache | 1 (3.1%) | 1 (3.1%) | 0 (0%) |
| Insomnia | 1 (3.1%) | 1 (3.1%) | 0 (0%) |
| Rash – pustular | 1 (3.1%) | 1 (3.1%) | 0 (0%) |

**Supplemental Table S5. Dose reductions/holds due to toxicity for each drug for the phase II portion of the clinical trial.**

| | **Number of Patients (%), Everolimus** | **Number of Patients (%), Exemestane** | **Number of Patients (%), Palbociclib** |
|---|---|---|---|
| **Dose was held** | 22 (68.8%) | 2 (6.3%) | 25 (78.1%) |
| **Dose reduction** | 13 (40.6%) | 0 (0%) | 16 (50.0%) |
| **Number of dose reductions** | | | |
| 1 dose | 13 (40.6%) | 0 (0%) | 15 (46.9%) |
| 2 doses | 0 (0%) | 0 (0%) | 1 (3.1%) |
